# A study of the benefits of vaccine mandates and vaccine passports for SARS-CoV-2

**DOI:** 10.1101/2021.11.10.21266188

**Authors:** Aaron Prosser, David L. Streiner

**Affiliations:** Department of Psychiatry and Behavioural Neurosciences, McMaster University

## Abstract

**Objective:** To evaluate the benefits of vaccine mandates and vaccine passports (VMVP) for SARS-CoV-2 by estimating the benefits of vaccination and exclusion of unvaccinated people from different settings.

**Methods:** Quantified the benefits of vaccination using meta-analyses of randomized controlled trials (RCTs), cohort studies, and transmission studies to estimate the relative risk reduction (RRR), absolute risk reduction (ARR), and number needed to vaccinate (NNV) for transmission, infection, and severe illness/hospitalization. Estimated the baseline infection risk and the baseline transmission risks for different settings. Quantified the benefits of exclusion using these data to estimate the number of unvaccinated people needed to exclude (NNE) to prevent one transmission in different settings. Modelled how the benefits of vaccination and exclusion change as a function of baseline infection risk. Studies were identified from recent systematic reviews and a search of MEDLINE, MEDLINE In-Process, Embase, Global Health, and Google Scholar.

**Results:** Data on infection and severe illness/hospitalization were obtained from 10 RCTs and 19 cohort studies of SARS-CoV-2 vaccines, totalling 5,575,049 vaccinated and 4,341,745 unvaccinated participants. Data from 7 transmission studies were obtained, totalling 557,020 index cases, 49,328 contacts of vaccinated index cases, and 1,294,372 contacts of unvaccinated index cases. The estimated baseline infection risk in the general population is 3.04%. The estimated breakthrough infection risk in the vaccinated population is 0.57%. Vaccines are very effective at reducing the risk of infection (RRR=88%, ARR=2.59%, NNV=39) and severe illness/hospitalization (RRR=89%, ARR=0.15%, NNV=676) in the general population. While the latter effect is small, vaccines nearly eliminate the baseline risk of severe illness/hospitalization (0.16%). Among an infected person’s closest contacts (primarily household members), vaccines reduce transmission risk (RRR=41%, ARR=11.04%, NNV=9). In the general population, the effect of vaccines on transmission risk is likely very small for most settings and baseline infection risks (NNVs ≥ 1,000). Infected vaccinated people have a nontrivial transmission risk for their closest contacts (14.35%), but it is less than unvaccinated people (23.91%). The transmission risk reduction gained by excluding unvaccinated people is very small for most settings: healthcare (NNE=4,699), work/study places (NNE=2,193), meals/gatherings (NNE=531), public places (NNE=1,731), daily conversation (NNE=587), and transportation (NNE=4,699). Exclusion starts showing benefits on transmission risk for some settings when the baseline infection risk is between 10% to 20%.

**Conclusions:** The benefits of VMVP are clear: the coercive element to these policies will likely lead to increased vaccination levels. Our study shows that higher vaccination levels will drive infections lower and almost eliminate severe illness/hospitalization from the general population. This will substantially lower the burden on healthcare systems. The benefits of exclusion are less clear. The NNEs suggest that hundreds, and even thousands, of unvaccinated people may need to be excluded from various settings to prevent one SARS-CoV-2 transmission from unvaccinated people. Therefore, consideration of the costs of exclusion is warranted, including staffing shortages from losing unvaccinated healthcare workers, unemployment/unemployability, financial hardship for unvaccinated people, and the creation of a class of citizens who are not allowed to fully participate in many areas of society.

**Registration:** This study is not registered.

**Funding:** This study received no grant from any funding agency, commercial, or not-for-profit sectors. It has also received no support of any kind from any individual or organization.

## Introduction

Vaccine mandates and vaccine passports (VMVP) for severe acute respiratory syndrome coronavirus 2 (SARS-CoV-2) are being implemented globally because their benefits may return societies to normality. These policies reduce risks (e.g., infection, transmission) by (1) increasing vaccination levels through coercion and (2) excluding those who remain unvaccinated from many areas (e.g., work, leisure, transportation). Increased vaccination moves populations closer to herd immunity, reducing the burden of SARS-CoV-2. Exclusion is meant to reduce the transmission risk posed by unvaccinated populations and is a key justification for VMVP.^1^ There are three uncertainties which make it hard to analyse the benefits of VMVP.

First, we do not know how effective VMVP will be at increasing vaccination and how many unvaccinated people need to be excluded to do so. Only controlled trials can determine this prospectively, given potential confounders driving any observed increases in vaccination. However, such trials are not feasible. Determining effectiveness retrospectively is also difficult because of confounders (e.g., differences in implementation, enforcement) and risks waiting for harm to be done before effectiveness is known. We can, however, analyse the benefits of vaccination and excluding unvaccinated people – the goal of this study – to better understand the benefits of VMVP.

Second, while vaccines clearly reduce many risks of SARS-CoV-2, the transmission data are only starting to emerge. Our study identified a growing literature on vaccines and transmission.^2–13^ These studies show benefit for transmission, but only report various metrics related to the relative risk reduction (RRR). RRR is 1 minus the relative risk (RR), where RR is the ratio of case rates in the vaccinated vs. unvaccinated groups. The absolute risk reduction (ARR) is another helpful way to quantify benefit using the risk difference between unvaccinated vs. vaccinated groups. It is the basis for the number needed to vaccinate (NNV=1/ARR), analogous to the number needed to treat (NNT). NNV is the number needed to vaccinate to prevent one case. Baseline risks influence the ARR and NNV, such that the lower the baseline risk, the lower the ARR and the higher the NNV, and vice versa. Baseline risks for SARS-CoV-2 are the cumulative incidence of the outcome in the unvaccinated general population. They are partly a property of the disease (e.g., virulence), time at risk, setting (e.g., proximity), and individuals (e.g., age). The RRR is more constant across baseline risks because it is mainly a property of treatments. Both the RRR and ARR are useful, but the latter is necessary to contextualize treatment benefit within the baseline risk. Thus, ARRs and NNVs are needed to quantify the absolute benefits of VMVP on transmission risk.

A third uncertainty is the number of unvaccinated people we need to exclude to reduce transmission risk. This is critical to determining the benefits of exclusion. Doing so requires using baseline transmission risks to calculate a metric similar to the NNV, which we call the ‘number needed to exclude’ (NNE=1/ARR). The difference is that the ARR of the NNE is the baseline transmission risk. The basis for the NNE is that exclusion removes *all* unvaccinated people, such that the ARR is just the baseline transmission risk in the unvaccinated population. Thus, the NNE is the number of unvaccinated people who must be excluded from a setting to prevent one transmission *from unvaccinated people* in that setting. NNE is helpful because it quantifies how many unvaccinated people need to be excluded to reduce the transmission risk *specific* to unvaccinated populations. The NNE does not tell us the number of unvaccinated people who need to be excluded by VMVP as the “price” for increased vaccinations leading to reduced transmissions overall. This depends on the NNE and NNV, in addition to vaccination levels in the general population in response to VMVP. For example, if VMVP increase vaccination levels to 90% in a population of 1,000 unvaccinated people, 100 people will be excluded. These 100 people are the “price” society pays for the transmission risk reductions driven by increased vaccinations in response to VMVP. This number needed to “pay” (NNP) for each transmission prevented overall can be estimated using the NNE and NNV and modelling different vaccination levels in the general population (see Methods).

Critically, societies need to be mindful of baseline transmission risks because when they are low, the NNE increases exponentially (Figure 1). Figure 1 shows that the transmission risk reduction gained by excluding unvaccinated people becomes very small when baseline risks are low, especially below 0.40% to 0.30%. This is the range of the ARRs of acetylsalicylic acid (ASA) in primary prevention of cardiovascular disease (CVD) outcomes (NNTs ≥250 to 333).^14–16^

**Figure 1.**
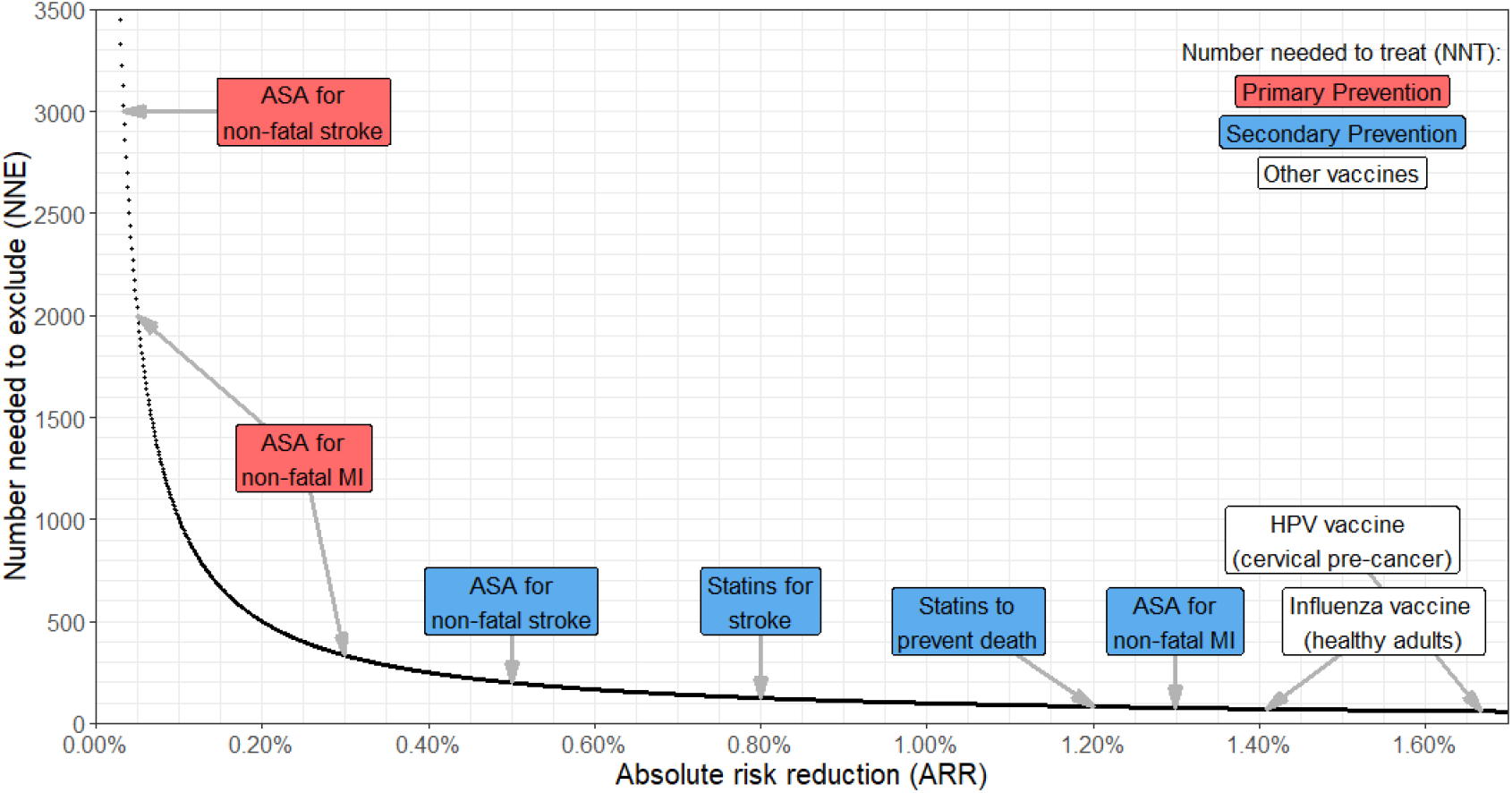
Relationship between baseline transmission risk and the number needed to exclude (NNE). The number needed to treat (NNT) has the same scale as the NNE (NNT=1/ARR). The only difference is that the ARR of the NNE is the baseline transmission risk. This figure shows that the risk reduction gained by excluding unvaccinated people becomes very small when baseline risks are low, especially when they are below 0.40% to 0.30%. This is the range of the ARRs of acetylsalicylic acid (ASA) in primary prevention of cardiovascular disease outcomes.^14–16^ The NNTs of other interventions are shown for comparison.^14 15 25 26 29 30^ ARR=absolute risk reduction. MI=myocardial infarction. HPV=human papillomaviruses.

The purpose of this study was to use meta-analyses to help address these uncertainties by estimating the benefits of VMVP in terms of vaccination and exclusion. First, we estimated the benefits of SARS-CoV-2 vaccines on reducing transmission, infection, severe illness/hospitalization, and death. We did this by conducting a meta-analysis of the transmission studies and another of the randomized controlled trials (RCTs) and cohort studies of SARS-CoV-2 vaccines identified in recent systematic reviews.^17–19^ The meta-analytic estimates of the RRR, ARR, and NNV for these outcomes allowed us to quantify the relative and absolute benefits of VMVP-driven increases in vaccination. Second, we quantified the absolute benefits of excluding unvaccinated people by calculating the NNE and NNP from our estimates of (1) the baseline infection risk in the general population and (2) the baseline transmission risks for different settings (e.g., household, healthcare, work). We also modelled how the NNE and NNP change if baseline infection risks increase as societies reopen. Finally, we offer indicators societies can use to determine when to relax VMVP.

## Methods

### Selection of studies and search criteria

We selected RCTs and cohort studies of adults identified in three recent systematic reviews.^17–19^ Transmission risks and their 95% confidence intervals (CIs) were extracted from a recent systematic review and meta-analysis by Zhao et al (2021)^20^ of the higher-quality studies. Zhao et al reported the transmission risks for different settings: households, healthcare, work/study places, meals/gatherings, public places, daily conversation, and transportation. These studies were all pre-vaccination and thus the estimates were from unvaccinated individuals. We performed a systematic review and meta-analysis of the studies examining the effect of SARS-CoV-2 vaccines on transmission. We searched MEDLINE, MEDLINE In-Process, Embase, and Global Health from the date of their inception to October 29, 2021, with no language restrictions. We used the search terms “coronavirus disease 2019”, “COVID-19”, “COVID”, “severe acute respiratory syndrome coronavirus 2”, or “SARS-CoV-2”, combined with “vaccine*”, combined with “transmission*”, “secondary attack*”, “SAR”, “contact trac*”, or “close contact*”. We identified additional reports by searching the reference lists and using Google Scholar’s cited-by function of the included studies.

### Outcomes and extraction

Our primary outcomes were the number of cases of transmission, infection, severe illness/hospitalization, and SARS-CoV-2-related death for vaccinated vs. unvaccinated groups and their sample sizes. Using the total sample sizes in the denominators facilitated estimation of the general population effects. Secondary outcomes were the number of cases of severe illness/hospitalization and SARS-CoV-2-related death among infected persons for vaccinated vs. unvaccinated groups. The denominators were the number of infections in each group. Details are in Supplementary Table S1. Data on fully vaccinated groups was extracted whenever possible. Total sample sizes were approximated from the reports of two cohort studies, and we used the number of tests in two other cohort studies. All but two studies reported infection data. If multiple metrics of infection were reported (e.g., all vs. symptomatic vs. asymptomatic), the higher number was extracted. If multiple metrics of severe illness/hospitalization were reported, the higher number was extracted. If reported, we extracted data on the groups which were seronegative at baseline to minimize the confounding influence of natural immunity. Safety data were not extracted because this has been reviewed elsewhere^21^ showing that SARS-CoV-2 vaccines are safe in adults. One author (AP) performed the study selection and data extraction, which was twice checked to confirm reliability and fidelity. Raw data is freely available online: https://tinyurl.com/tvheppj4. Data were extracted from reports into standardized electronic forms. For the RCTs and cohort studies, data on vaccine, number of doses, sex, age, country, comparator, study period, and start of follow-up relative to dose were extracted. For the transmission studies, data on vaccine, number of doses, country, index case sample size, contact sample size, contact population, study period, and variant were extracted. Results are tabulated in the online supplement.

### Data analysis

For each study we calculated the RR, RRR, ARR, and NNV for the primary and secondary outcomes, in addition to the baseline risk (risk of the outcome in the unvaccinated group) and breakthrough risk (risk of the outcome in the vaccinated group). For the transmission studies we calculated the transmission risks for the vaccinated vs. index cases. Analyses were conducted in R with the *metafor* package.^22 23^ We used random effects (restricted maximum-likelihood estimator, REML) models to combine the log RRs and ARRs. The summary estimates were converted to the RRR and NNV. This allowed us to estimate the relative and absolute benefits of vaccines on reducing transmission, infection, severe illness/hospitalization, and death in the general population and among infected persons. The *I*^2^ statistic and its 95% CI were calculated to convey the amount of heterogeneity.

We used random effects (REML) models to estimate the baseline, breakthrough, and transmission risks. The most important were the baseline infection risk and the unvaccinated transmission risk. Note, unvaccinated transmission risks are not true baseline risks. This is because they are the risk among infected unvaccinated index cases, not the unvaccinated general population per se. Estimating baseline transmission risks requires calculating the combined probability of infection *and* transmission because a person must be infected first before they can transmit SARS-CoV-2. Thus, baseline transmission risks for different settings were estimated by multiplying the unvaccinated transmission risks in Zhao et al and our analysis by the baseline infection risk (3.04%) we estimated from the RCTs and cohort studies (Supplementary Table S3). We modelled the effect of increasing baseline infection risks (5%, 10%, and 20%) on baseline transmission risks to simulate different scenarios as societies reopen. 20% is a “worst case scenario” since the baseline infection risk would be in the range of the unvaccinated transmission risk (23.91%) among an infected person’s closest contacts estimated in our analysis (Supplementary Table S8).

The baseline transmission risks and the RRR of vaccines on transmission allowed us to calculate four key metrics for each setting and baseline infection risk (3.04%, 5%, 10%, 20%). The first was the NNE. The second was the ARR and NNV of vaccines on transmission risk. ARRs were calculated by multiplying baseline transmission risks (Table 2) by the RRR we estimated from the large transmission studies in our meta-analysis (0.41, 41%) (Supplementary Table S8). Recall, the ARR is the raw difference between the risks in the unvaccinated vs. vaccinated groups and RRR is the percent difference between them (1 minus RR). Since we know the baseline transmission risks (Table 2), multiplying each risk by the RRR gives the ARR and thus the NNV. One can apply the same RRR across baseline risks to estimate the ARRs and NNVs because RRRs are fairly constant across baseline risks since they are a property of vaccines.

The third metric was the vaccinated transmission risk. These were estimated by subtracting the ARR (Table 3) from its corresponding baseline transmission risk (Table 2). The residual is the transmission risk the vaccinated population poses to others.

The fourth metric was the NNP. The NNP is calculated:

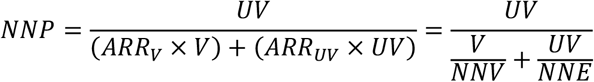

*ARR*_*V*_ is the ARR of vaccines on transmission in that setting (Supplementary Table S10) and *ARR*_*UV*_ is the ARR of exclusion on transmission in that setting (i.e., the baseline transmission risk) (Table 2). The ARRs are weighted by the effectiveness of VMVP (i.e., vaccination levels in the general population). *V* is the proportion of people in the general population who get vaccinated. *UV* is the proportion of people in the general population who remain unvaccinated (*UV* = 1 − *V*). The numerator is the total number (expressed as a proportion) of unvaccinated people at a given vaccination level. The denominator is the overall ARR of VMVP on transmission in a setting at a given vaccination level. The denominator can be expressed equivalently in terms of the NNV and NNE, which expresses the total number of transmissions prevented by VMVP in a setting at a given vaccination level.

The NNP tells us—at a given level of vaccination in the general population—how many unvaccinated people need to be excluded from a setting for every one transmission prevented overall in that setting (from exclusion *and* vaccination). Thus, the value of the NNP says that, for every one transmission prevented overall, *X* number of unvaccinated people need to be excluded. The NNP is the “price” society pays for preventing one transmission overall. However, the NNP does *not* tell us how many unvaccinated people need to be excluded to reduce the risks specific to unvaccinated populations. The NNE quantifies this. The NNE tells us how many unvaccinated people must be excluded from a setting to prevent one transmission *from unvaccinated people* in that setting, regardless of the level of vaccination in the general population. If vaccination levels are 100%, then the NNP equals zero because *UV* equals zero (there are no unvaccinated people to exclude). If vaccination levels are 0%, NNP reduces to the NNE because *V* equals zero (everyone must be excluded because everyone is unvaccinated). The NNP is always less than the NNE (unless *V* equals zero) because of the additional risk reduction gained from vaccinations.

## Results

10 RCTs and 19 cohort studies of SARS-CoV-2 vaccines on infection, severe illness/hospitalization, and death were identified in the systematic reviews^17–19^ and included in our meta-analysis (Supplementary Table S1). Excluded reports are in Supplementary Table S2. 5,575,049 vaccinated (V) and 4,341,745 unvaccinated (UV) participants were included in our analysis of infection, which was the largest analysis (Supplementary Table S3). The vaccines included BNT162b2 (Comirnaty, BioNTech/Pfizer: *n* effects=14, V=5,337,467, UV=4,202,165), ChAdOx1 nCoV-19 (Vaxzevria, AstraZeneca: *n* effects=6, V=96,745, UV=813,192), Ad26.COV2.S (Janssen: *n* effects=2, V=21,293, UV=37,288), mRNA-1273 (Spikevax, Moderna: *n* effects=2, V=17,514, UV=28,770), WIV04 (Sinopharm: *n* effects=1, V=12,743, UV=12,737), HB02 (Sinopharm: *n* effects=1, V=12,726, UV=12,737), Gam-COVID-Vac (Sputnik V: *n* effects=1, V=14,964, UV=4,902), and NVX-CoV2373 (Novavax: *n* effects=1, V=1,357, UV=1,327). There were four reports of multiple vaccines (mainly BNT162b2 and ChAdOx1 nCoV-19) which did not report the effects separately and were analysed as a group. The sizes of the other primary and secondary outcome analyses are in Supplementary Tables S4-7.

7,059 citations were identified by the database search and 6 from the reference list/Google scholar cited-by function search (Supplementary Figure S1). 29 potentially eligible articles were retrieved in full-text. 7 studies of the effect of vaccines on transmission were included in our meta-analysis (Supplementary Table S8). Excluded reports are in Supplementary Table S9. This is a rapidly emerging literature since most reports were posted in August to October 2021. Across these 7 studies there were 49,328 contacts of vaccinated index cases and 1,294,372 contacts of unvaccinated index cases. These were the closest contacts of the index cases, primarily household members. There was a total of 557,020 index cases. There were two small studies with less than 300 index cases.^5 7^ We describe the meta-analysis of the large studies to provide more robust estimates. Study periods were between December 2020 to September 2021. Studies were from the Netherlands,^2 13^ UK,^3 4 7^ Israel,^5^ and Spain.^6^ All but one study^4^ reported the dominant variant: Alpha (*n*=2),^2 5^ Delta (*n*=2),^7 13^ and ∼50% Alpha and ∼50% delta (*n*=2).^3 6^

Table 1 summarizes the meta-analysis of the baseline risks and the effect of vaccines on risk of infection, severe illness/hospitalization, and death (see Supplementary Tables S3-7 for details). The estimated baseline infection risk in the general population is 3.04%. This is consistent with the very crude baseline infection risk (3.15%) using the number of confirmed SARS-CoV-2 globally (248,467,363 as of 5-Nov-2021) and the global population (7.9 billion).^24^ The estimated breakthrough infection risk in the vaccinated population is 0.57%. The estimated baseline risk of severe illness/hospitalization in the general population is 0.16%. Among infected persons, it is 11.02%. This indicates that SARS-CoV-2 is a disease with a high risk of morbidity for people in their middle ages or older, which was the age range of most participants in the RCTs and cohort studies (Supplementary Table S1). There were too few SARS-CoV-2-related deaths in the studies to meaningfully analyse these data.

**Table 1.** Meta-analysis of SARS-CoV-2 vaccines on infection, severe illness/hospitalization, and death

|  |  |  |  |  |
| --- | --- | --- | --- | --- |
| Outcome | Baseline risk†<br>(95% CI) | Effect of vaccination‡ |  |  |
|  |  | RRR<br>(95% CI) | ARR<br>(95% CI) | NNV<br>(95% CI) |
| General population |  |  |  |  |
| Infection | 3.04%<br>(2.08%, 4.01%) | 88%<br>(83%, 92%) | 2.59%<br>(1.75%, 3.43%) | 39<br>(29, 57) |
| Severe illness/hospitalization | 0.16%<br>(0.09%, 0.23%) | 89%<br>(82%, 93%) | 0.15%<br>(0.09%, 0.20%) | 676<br>(489, 1,094) |
| Death | Insufficient data | Insufficient data | Insufficient data | Insufficient data |
| Among infected persons |  |  |  |  |
| Severe illness/hospitalization§ | 11.02%<br>(6.31%, 15.72%) | 7%<br>(-55%, 44%) | 5.64%<br>(0.69%, 10.59%) | 18<br>(9, 146) |
| Death | Insufficient data | Insufficient data | Insufficient data | Insufficient data |

RRR=relative risk reduction (1 – relative risk). ARR=absolute risk reduction. NNV=number needed to vaccinate. 95% CI=95% confidence interval. Brackets are the 95% confidence intervals (lower limit, upper limit).
<sup>‡</sup>See Tables S3-7 for details.
<sup>†</sup>Baseline risk is defined as the risk of the SARS-CoV-2 outcome (e.g., infection, severe illness/hospitalization, death) in the unvaccinated population.
§Statistically underpowered due to the small numbers of infections in the vaccinated groups for most studies. The RRR, ARR, and NNV are not precise estimates.

SARS-CoV-2 vaccines are very effective at reducing the risk of infection (RRR=88%, ARR=2.59%, NNV=39) and severe illness/hospitalization (RRR=89%, ARR=0.15%, NNV=676) in the general population. While the latter effect is small, the ARR is almost the same as the baseline risk (0.16%). This means vaccines almost eliminate the baseline risk of severe illness/hospitalization in the general population. The secondary analysis of severe illness/hospitalization among infected persons was statistically underpowered due to the small numbers of infections among the vaccinated. However, if a person is infected, vaccines partially protect against severe illness/hospitalization (ARR=5.64%). Among an infected person’s closest contacts (primarily household members), vaccines reduce transmission risk, but the RRR is smaller than for infection (RRR=41%, ARR=11.04%, NNV=9) (Supplementary Table S8). The ARR is high because the unvaccinated transmission risk is 23.91% among their closest contacts.

Supplementary Table S10 summarizes the estimated effects of SARS-CoV-2 vaccines on transmission risk among infected persons and in the general population for different settings. In the general population, the effects are very small for most settings and baseline infection risks (NNVs ≥ 1,000). The reason is because, while vaccines reduce transmission risk (41%), baseline transmission risks are very low to begin with. Supplementary Table S11 summarizes the transmission risks of vaccinated people in different settings. The transmission risk post-vaccination among infected persons is notable for some settings: closest contacts (14.35%), households (7.08%), meals/gatherings (3.66%), and daily conversation (3.30%). This means there is a nontrivial transmission risk if a vaccinated person is infected, especially for their closest contacts.

Table 2 summarizes the estimated benefits of excluding unvaccinated people on transmission risk among infected persons and in the general population. For a baseline infection risk of 3.04%, the transmission risk reduction in the general population gained by excluding unvaccinated people is very small for most settings: healthcare (NNE=4,699), work/study places (NNE=2,193), meals/gatherings (NNE=531), public places (NNE=1,731), daily conversation (NNE=587), and transportation (NNE=4,699). These NNEs are within and some are above the NNTs of ASA for primary prevention in CVD (NNTs ≥250).^14–16^ The NNEs are similar if the baseline infection risk is 5%. Exclusion starts showing benefits for some settings between 10% to 20% baseline infection risk: meals/gatherings (NNE=161 to 81), public places (NNE=526 to 263), and daily conversation (NNE=179 to 89). However, the benefits of exclusion remain small for healthcare (NNE=714), work/study places (NNE=333), and transportation (NNE=714) even if the baseline infection risk is 20%.

**Table 2.**
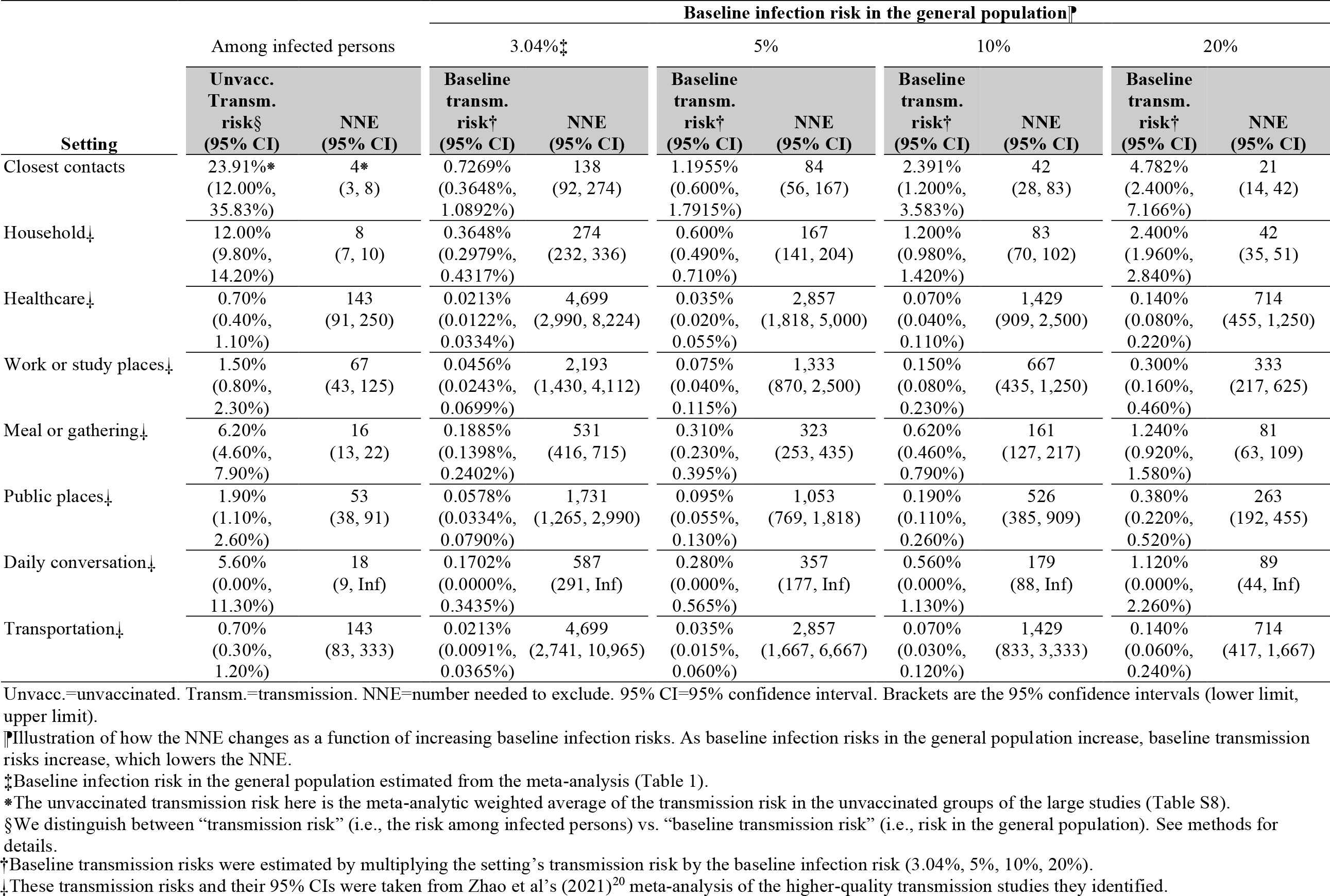
Estimated effects of excluding unvaccinated people on transmission risk among infected persons and in the general population

Figure 2 plots the NNPs for different settings, baseline infection risks, and vaccination levels. The 90-100% ranges are shown to illustrate the effects of high levels of vaccination in the general population. For a baseline infection risk of 3.04%, the numbers of unvaccinated people excluded for every one transmission prevented overall starts to be relatively few (NNP < 250) in daily conversations and meals/gatherings at ∼75% vaccination. Public places and work/study places only reach this point at ∼95% vaccination, whereas healthcare and transportation start at ∼98% vaccination. The NNP remains over 1,000 for healthcare and transportation up until ∼90% vaccination. If baseline infection risks increase to 10% or 20%, the NNPs decline substantially because the NNVs and NNEs decrease (Supplementary Table S10 and Table 2). This means that, when baseline infection risks are high, fewer unvaccinated people need to be excluded for every one transmission prevented overall by VMVP.

**Figure 2.**
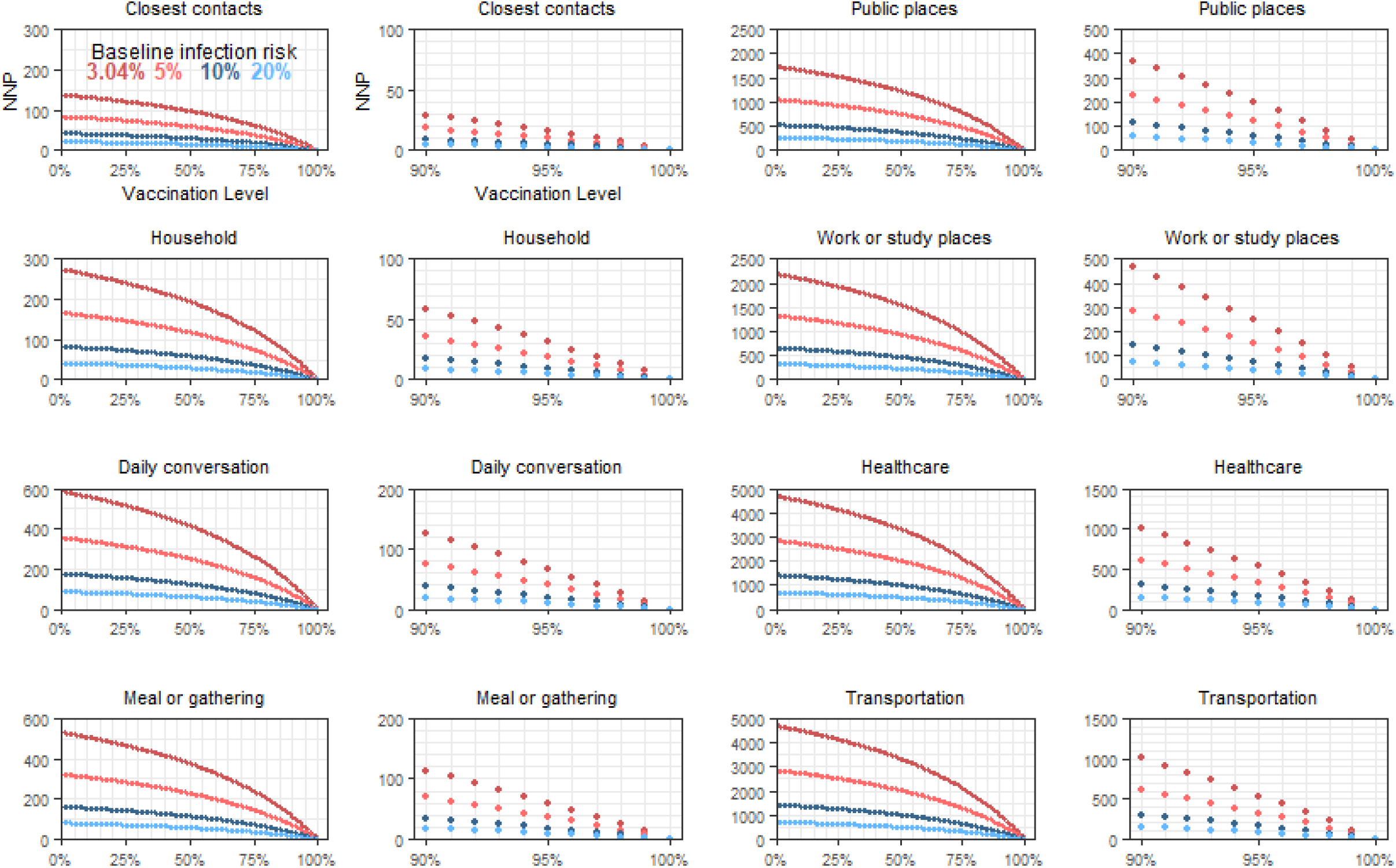
Modelling the number needed to pay (NNP) in terms of excluding unvaccinated people to prevent one transmission overall in different settings as a function of baseline infection risk and vaccination levels in the general population.

## Discussion

VMVP use coercion to increase vaccination levels and exclude those who remain unvaccinated from many areas (e.g., work, leisure, transportation). Exclusion is meant to reduce the transmission risk posed by unvaccinated populations and is a key justification for VMVP.^1^ We ran large-scale meta-analyses of SARS-CoV-2 vaccines which included hundreds of thousands of participants to estimate the benefits of these two aspects of VMVP. We found that SARS-CoV-2 vaccines are very effective at reducing the relative and absolute risk of infection (RRR=88%, ARR=2.59%, NNV=39) and severe illness/hospitalization (RRR=89%, ARR=0.15%, NNV=676) in the general population. While the latter effect is small, vaccines nearly eliminate the baseline risk of severe illness/hospitalization (0.16%). The NNV for infection is similar to influenza and HPV vaccines.^25 26^ Among an infected person’s closest contacts (primarily household members), vaccines reduce transmission risk, but the RRR is smaller than for infection (RRR=41%, ARR=11.04%, NNV=9).

The benefits of excluding unvaccinated people on reducing transmission risk are less clear. We estimated the baseline infection risk (3.04%) from the analysed studies to calculate the baseline transmission risk in the general population for different settings. From these we could calculate the NNE and NNP, which quantify different aspects of the risk reductions gained from excluding unvaccinated people. We also modelled how the NNE and NNP change depending on the baseline infection risk, including a “worst case scenario” of a 20% baseline risk. The NNEs show that the benefits of exclusion are in the range (and sometimes higher) than the NNTs of ASA for primary prevention in CVD.^14–16^ This is notable given that ASA is not generally recommended for primary prevention in all adults because the very small benefits (NNTs ≥250 to 333) do not outweigh the risk of harm.^16^ The estimated NNEs suggest that hundreds, and even thousands, of unvaccinated people may need to be excluded from various settings to prevent one SARS-CoV-2 transmission from unvaccinated people (Table 2). Exclusion starts showing benefits on transmission risk for some settings when the baseline infection risk is between 10% to 20%.

The NNPs (Figure 2) suggest that, at a baseline infection risk of 3.04%, vaccination levels may need to reach at least ∼95% to ∼98% before only relatively few unvaccinated people (NNP < 250) are excluded for every one transmission prevented overall in public areas, work/study places, healthcare, and transportation. They may need to reach at least ∼75% before the same effects are seen in daily conversations and meals/gatherings. Until ∼90% vaccination, more than 1,000 unvaccinated people may need to be excluded to prevent one transmission overall in healthcare and transportation settings. Since the NNPs are less than the NNEs, excluding, for instance, 1,000 unvaccinated people from healthcare settings still does not prevent one transmission *from unvaccinated people* since the NNE for healthcare is 4,699 (at a 3.04% baseline infection risk). Rather, excluding these 1,000 people is the “price” society pays for the benefits of 90% vaccination, but excluding them falls short of reducing the transmission risk specific to those unvaccinated people. Finally, we found that infected vaccinated people have nontrivial transmission risks for their closest contacts (14.35%). This is consistent with emerging epidemiological data showing no correlation between the level of full vaccination and the incidence of new SARS-CoV-2 infections.^27^

The benefits of VMVP are clear: the coercive element to these policies will likely lead to increased vaccination levels. Our study shows that higher vaccination levels will drive infections lower and almost eliminate severe illness/hospitalization from the general population. This will substantially lower the burden on healthcare systems. It also explains why unvaccinated patients now constitute most SARS-CoV-2 hospitalizations and intensive care unit (ICU) admissions.^28^ Among infected persons, vaccines reduce the risk of transmission to their closest contacts by about 41%. However, in the general population, the magnitude of this effect is likely very small (in the range of ASA in primary prevention of CVD).

These benefits come with costs. An unknown percentage of people will not get vaccinated despite VMVP. This means those unvaccinated people will be excluded from many areas of modern life (e.g., many places of work, leisure, transportation, education), especially work in healthcare settings. The NNEs and NNPs show that the transmission risk reduction gained by excluding unvaccinated people is very small for most settings (especially healthcare and transportation settings). In light of this, there are at least three costs to consider. The first is healthcare-specific. There is a possibility of staffing shortages from the loss of unvaccinated workers, which may have downstream effects on patient care. This may be limited by increases in vaccinations in response to VMVP. Second, there is the cost of unemployment and – in some cases – unemployability if vaccines are a condition of employment or higher-education in most organizations. This would mean unvaccinated individuals and their families will face financial hardship. This may have downstream effects on their mental/physical health. A third cost of exclusion is the creation of a class of citizens who are not allowed to fully participate in many areas of society.

Baseline infection risk is critically important to evaluating the costs vs. benefits of excluding unvaccinated people. Therefore, we suggest that baseline infection risk be included as a key indicator for when to relax VMVP. It compliments other indicators, such as hospital/ICU occupancy and the basic and effective reproduction number. As societies reopen, the baseline infection risk may increase. However, it is also possible that the baseline infection risk will not increase dramatically due to high levels of vaccination and natural immunity which likely already exist in many societies. If baseline infection risks remain or decline below 3.04% (3,040 cases per 100,000) over a 1-to-3 month period (the typical follow-up period of the studies in our meta-analysis), then consideration of whether the costs of exclusion outweigh the benefits is warranted.

Our study has some limitations. Our estimates derive from studies during a period of the pandemic when physical distancing, lockdowns, and universal masking were in place, which contained infection rates. We have tried to address this by modelling how the NNE and NNP change with increased baseline infections. We were unable to answer how much VMVP will increase vaccination levels, which requires controlled trials and cohort studies. There were too few transmission studies to perform a meta-regression/subgroup analysis or examine the moderating effects of variants and vaccines. Not all SARS-CoV-2 vaccines were included in our meta-analysis and BNT162b2 and ChAdOx1 nCoV-19 were by far the most studied. Our conclusions do not generalize to child/adolescent populations. We were unable to analyse the impact of natural immunity on reducing transmission risk. In addition to more studies of vaccines on transmission, future research should focus on health economic analyses of the costs vs. benefits of exclusion, natural immunity and transmission, the impact of variants, and quantifying what is an acceptable baseline risk of infection and transmission in terms of health and economic outcomes.

Notwithstanding these limitations, our study has strengths. Our estimates of the effect of SARS-CoV-2 vaccines on transmission, infection, and severe illness/hospitalization in the general population are robust because they are based on hundreds of thousands of participants. Using a recent meta-analysis^20^ also provided data for robust estimates of the baseline transmission risks used to calculate the NNEs and NNPs for different settings. We performed the first meta-analysis of the studies of vaccines on transmission, a rapidly emerging literature. Our study is also the first to quantify the benefits of exclusion on transmission risk. We hope that these results will assist societies and policymakers in their cost vs. benefit analyses of VMVP.

## Supporting information

dat_covidVMVP

dat_Zhao_etal

## Data Availability

Raw data is freely available online: https://tinyurl.com/tvheppj4.

## Acknowledgement

We thank numerous colleagues who provided helpful comments and criticisms during the writing of this study. We are fully vaccinated healthcare workers.

## Contributions

AP and DS conceptualized the study, wrote/edited the manuscript, and had full access to the data. AP performed the study selection, data extraction, and data analysis. AP and DS conceptualized and developed the estimation and modelling procedures. AP is the corresponding author. All authors approved the final version of the manuscript. AP is the guarantor and attests that all listed authors meet authorship criteria and that no others meeting the criteria have been omitted.

AP affirms that this manuscript is an honest, accurate, and transparent account of the study being reported, that no important aspects of the study have been omitted, and that any discrepancies from the study as planned have been explained.

## Completing Interests

All authors have completed the ICMJE uniform disclosure form at http://www.icmje.org/disclosure-of-interest/ and declare: no support from any organisation for the submitted work; no financial relationships with any organisations that might have an interest in the submitted work in the previous three years; no other relationships or activities that could appear to have influenced the submitted work.

## Ethical approval

Not required.

## Data sharing

Raw data is freely available online: https://tinyurl.com/tvheppj4.

