## Supplementary material for "A study of the benefits of vaccine mandates and vaccine passports for SARS-CoV-2": Online Supplement

Figure S1. Prisma flow chart of the studies of SARS-CoV-2 vaccines on transmission risk

Table S1. Characteristics of the included RCTs and cohort studies of SARS-CoV-2 vaccines identified in recent systematic reviews[1-3]

Table S2. Excluded studies identified in recent systematic reviews

Table S3. Meta-analysis of SARS-CoV-2 vaccines on infection in the general population

Table S4. Meta-analysis of SARS-CoV-2 vaccines on severe illness/hospitalization in the general population

Table S5. Meta-analysis of SARS-CoV-2 vaccines on death in the general population

Table S6. Meta-analysis of SARS-CoV-2 vaccines on severe illness/hospitalization among infected persons

Table S7. Meta-analysis of SARS-CoV-2 vaccines on death among infected persons

Table S8. Characteristics and meta-analysis of studies of SARS-CoV-2 vaccines on transmission risk among infected persons

Table S9. Excluded studies identified in the systematic review of the studies of SARS-CoV-2 vaccines on transmission risk

Table S10. Estimated effects of SARS-CoV-2 vaccines on transmission risk among infected persons and in the general population

Table S11. Estimated transmission risks of vaccinated people among infected persons and in the general population

Table S12. PRISMA 2020 Checklist

**Figure S1.** PRISMA flow chart of the studies of SARS-CoV-2 vaccines on transmission risk

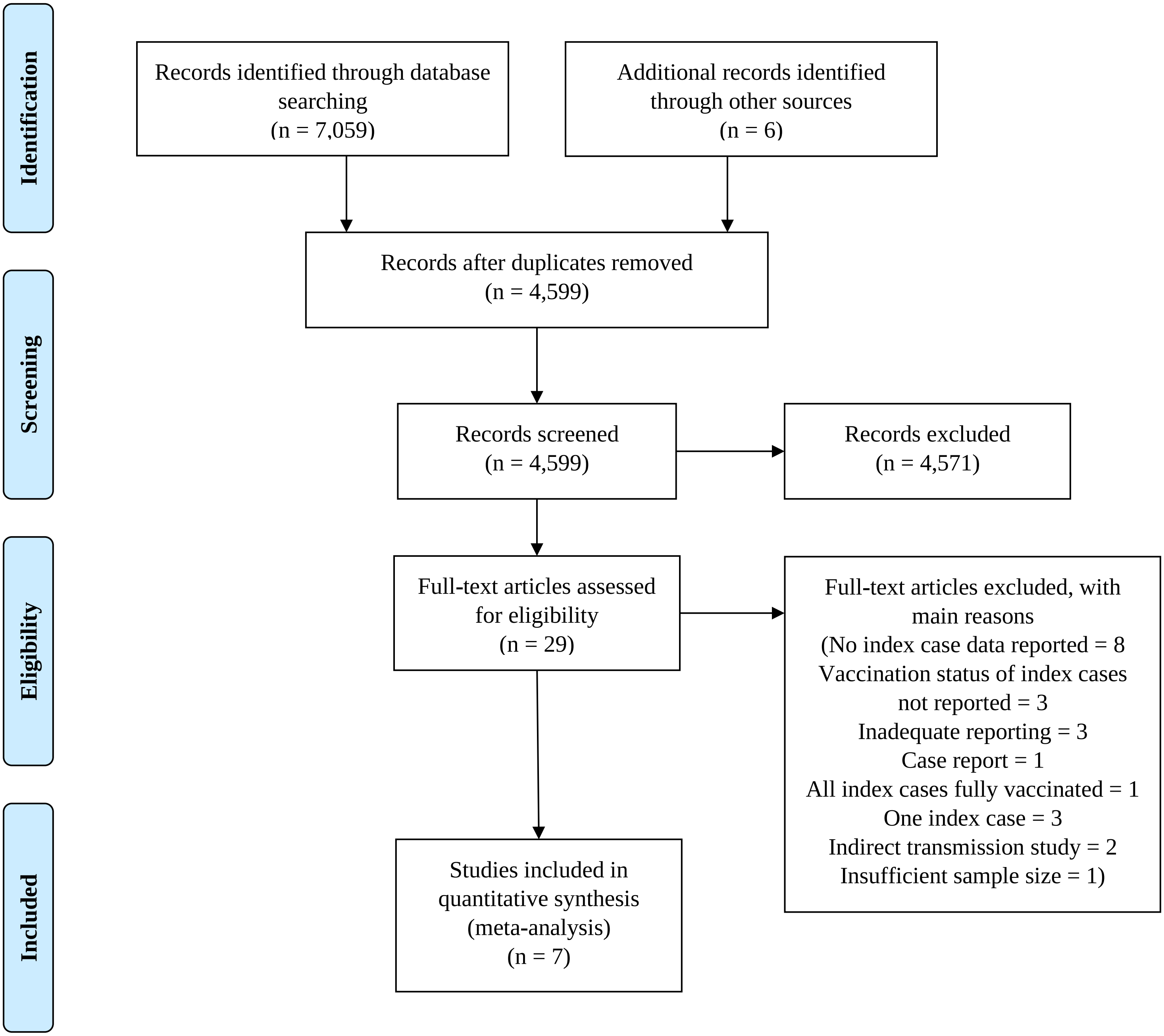

**Table S1.** Characteristics of the included RCTs and cohort studies of SARS-CoV-2 vaccines identified in recent systematic reviews[1-3]

| Study | Vaccine (doses) | Population | Study design | Sample size | Study period | Start of follow-up | Extracted SARS-CoV-2 outcomes |
|---|---|---|---|---|---|---|---|
| Amit et al (2021)[4] | BNT162b2 (at least 1 dose, 84% fully vaccinated) | *% female:* Not reported.<br>*Age:* Not reported.<br>*Country:* Israel | *Type:* Cohort<br>*Comparison:* Unvaccinated controls. | *Vaccinated:* 7,214†<br>*Unvaccinated:* 1,895† | 19-Dec-2020 to 24-Jan-2021 | 15 to 28 days post-first dose. | Infection |
| Björk et al (2021)[5] | BNT162b2 (at least 1 dose) | *% female:* 52.9%<br>*Age:* median 44 (range of total sample: 18 to 64).<br>*Country:* Sweden | *Type:* Cohort<br>*Comparison:* Unvaccinated controls. | *Vaccinated:* 26,587<br>*Unvaccinated:* 779,154 | 27-Dec-2020 to 28-Feb-2021 | 14-days post-first dose. | Infection<br>Death<br><br>*Authors report the case counts of the fully vaccinated group, but not the sample size. Thus, we could only extract data for those who received at least one dose.* |
| Dagan et al (2021)[6] | BNT162b2 (2 doses) | *% female:* 50%<br>*Age:* median 45 (IQR: 35 to 62)<br>*Country:* Israel | *Type:* Cohort<br>*Comparison:* Matched unvaccinated controls. | *Vaccinated∗:*<br>Infection – 108,529<br>Hospital – 110,054<br>Death – 110,101<br>*Unvaccinated∗:*<br>Infection – 107,209<br>Hospital – 109,867<br>Death – 110,008 | 20-Dec-2020 to 1-Feb-2021 | 7-days post-second dose. | Infection<br>Hospitalization<br>Death |
| Fabiani et al (2021)[7] | BNT162b2 (2 doses) | *% female:* 77.6%<br>*Age:* mean 47.1<br>*Country:* Italy | *Type:* Cohort<br>*Comparison:* Unvaccinated controls. | *Vaccinated:* 5,186<br>*Unvaccinated:* 1,090 | 27-Dec-2021 to 24-Mar-2021 | 7-days post-second dose. | Infection |
| Guijarro et al (2021)[8] | BNT162b2 (2 doses) | *% female:* Not reported.<br>*Age:* Not reported.<br>*Country:* Spain | *Type:* Cohort<br>*Comparison:* Unvaccinated controls. | *Vaccinated:* 2,116††<br>*Unvaccinated:* 170,513 | 21-Dec-2020 to 24-Feb-2021 | 7-days post-second dose. | Infection |
| Haas et al (2021)[9] | BNT162b2 (2 doses) | *% female:* 71.9% in the vaccinated group.<br>*Age:* 16 to ≥ 65. Full range, means, or medians not reported<br>*Country:* Israel | *Type:* Cohort<br>*Comparison:* Unvaccinated population. | *Vaccinated:* 4,714,932<br>*Unvaccinated:* 1,823,979∗∗ | 24-Jan-2021 to 3-April-2021 | 7-days post-second dose. | Infection<br>Hospitalization<br>Death |
| Jones et al (2021)[10] | BNT162b2 (1 dose) | *% female:* Not reported.<br>*Age:* Not reported.<br>*Country:* UK | *Type:* Cohort<br>*Comparison:* Unvaccinated controls | *Vaccinated:* 1,989§§<br>*Unvaccinated:* 3,252§§ | 18-Jan-2021 to 31-Jan-2021 | 12-days post-first dose. | Infection |

| | | | | | | | |
|---|---|---|---|---|---|---|---|
| Monge et al (2021)[11] | 99.8% were BNT162b2 (2 doses) | *% female:* 70.9%<br>*Age:* mean 85.9<br>*Country:* Spain | *Type:* Cohort<br>*Comparison:* Unvaccinated controls. | *Vaccinated:* 207,774<br>*Unvaccinated:* 230,438 | 27-Dec-2020 to 10-Mar-2021 | 29-days post-first dose. | Infection |
| Moustsen-Helms et al (2021)[12] | BNT162b2 (2 doses) | *% female:* 63.6% (long-term care facility patients) and 82.2% (healthcare workers)<br>*Age:* median 84 (IQR: 77 to 90, long-term care facility patients) and median 47 (IQR: 36 to 57, healthcare workers)<br>*Country:* Denmark | *Type:* Cohort<br>*Comparison:* Unvaccinated control. | *Vaccinated:* 114,406<br>*Unvaccinated:* 255,673<br><br>*Sample sizes are the combined totals of long-term care facility patients and healthcare workers.* | 27-Dec-2020 to 18-Feb-2021 | 7-days post-second dose. | Infection<br><br>*Authors reported long-term care facility patients and healthcare workers separately. We combined the case counts to increase the sample size.* |
| Polack et al (2020)[13] | BNT162b2 (2 doses) | *% female:* 49.4%<br>*Age:* median 52 (range: 16-91)<br>*Country:* United States, Argentina, Brazil, South Africa, Germany, Turkey. | *Type:* RCT<br>*Comparison:* Saline placebo. | *Vaccinated:* 18,198<br>*Unvaccinated:* 18,325 | 27-July-2020 to 14-Nov-2020 | 7-days post-second dose. | Infection<br>Severe illness<br>Death |
| Tang et al (2021)[14] | BNT162b2 (2 doses) | *% female:* 66% (vaccinated) and 58.3% (unvaccinated)<br>*Age:* 88.7% ≤ 65 (vaccinated) and 84.8% ≤ 65 (unvaccinated). Full range, means, or medians not reported.<br>*Country:* United States | *Type:* Cohort<br>*Comparison:* Unvaccinated controls. | *Vaccinated:* 2,724<br>*Unvaccinated:* 2,165 | 17-Dec-2020 to 20-Mar-2021 | 7-days post-second dose. | Infection |
| Khan & Mahmud (2021)[15] | BNT162b2 (2 doses) and mRNA-1273 (2 doses) | *% female:* 7.8%<br>*Age:* median 68<br>*Country:* United States<br><br>*All patients had inflammatory bowel disease. Many were on immunosuppressive medications.* | *Type:* Cohort<br>*Comparison:* Unvaccinated controls. | *BNT162b2:* 2,873<br>*mRNA-1273:* 3,380<br>*Unvaccinated:* 14,697 | 18-Dec-2020 to 20-Apr-2021 | 7-days post-second dose. | Infection<br>Severe illness<br><br>*Authors report all-cause mortality but not SARS-CoV-2-related death. They defined severe illness as either hospitalization or death but did not report death data separately.* |

| | | | | | | | |
|---|---|---|---|---|---|---|---|
| Swift et al (2021)[16] | BNT162b2 (2 doses) and mRNA-1273 (2 doses) | *% female:* 70.1%<br>*Age:* median 41<br>*Country:* United States | *Type:* Cohort<br>*Comparison:* Unvaccinated controls. | *Vaccinated:* 44,011<br>*Unvaccinated:* 23,931 | 1-Jan-2021 to 31-Mar-2021 | 14-days post-second dose. | Infection<br>*Data not separated by vaccine received* |
| Glampson et al (2021)[17] | BNT162b2 (at least 1 dose) and ChAdOx1 nCoV-19 (at least 1 dose) | *% female:* 48.5%<br>*Age:* 16 to ≥ 80. Full range, means, or medians not reported.<br>*Country:* UK | *Type:* Cohort<br>*Comparison:* Unvaccinated control. | *Vaccinated:* 389,587<br>*Unvaccinated:* 1,794,352 | 8-Dec-2020 to 24-Feb-2021 | X-days post-second dose. | Death<br>*The authors did not report the case counts for infection or hospitalization in the unvaccinated group and did not separate the death data by vaccine type.* |
| Lumley et al (2021)[18] | BNT162b2 (2 doses) and ChAdOx1 nCoV-19 (2 doses) | *% female:* 74%<br>*Age:* median 39 (IQR: 30 to 50)<br>*Country:* UK | *Type:* Cohort<br>*Comparison:* Unvaccinated controls. | *Vaccinated:* 940<br>*Unvaccinated:* 10,513 | 1-Sept-2020 to 28-Feb-2021 | 14-days post-second dose. | Infection<br>Hospitalization<br>*The authors did not separate the cases by vaccine type.* |
| Menni et al (2021)[19] | BNT162b2 (1 dose) and ChAdOx1 nCoV-19 (1 dose) | *% female:* 59.5%<br>*Age:* mean 62.7<br>*Country:* UK | *Type:* Cohort<br>*Comparison:* Unvaccinated controls. | *BNT162b2:* 67,293<br>*ChAdOx1 nCoV-19:* 36,329<br>*Unvaccinated:* 464,356 | 8-Dec-2020 to 10-Mar-2021 | From the first dose. | Infection |
| Pritchard et al (2021)[20] | BNT162b2 (2 doses) and ChAdOx1 nCoV-19 (2 doses) | *% female:* 53.5%<br>*Age:* 55 (IQR: 40 to 68)<br>*Country:* UK | *Type:* Cohort<br>*Comparison:* Unvaccinated controls. | *BNT162b2:* 57,646<br>*ChAdOx1 nCoV-19:* 41,018<br>*Unvaccinated:* 329,419 | 1-Dec-2020 to 8-May-2021 | Post-second dose (21 days post-first dose). | Infection |
| Shrotri et al (2021)[21] | BNT162b2 (at least 1 dose) and ChAdOx1 nCoV-19 (at least 1 dose) | *% female:* 69.6%<br>*Age:* median 86 (IQR: 80 to 91)<br>*Country:* UK | *Type:* Cohort<br>*Comparison:* Unvaccinated controls. | *Vaccinated:* 15,289‡‡<br>*Unvaccinated:* 15,392‡‡ | 11-June-2020 to 15-Mar-2021 | 14-days post-first dose. | Infection |
| Vasileiou et al (2021)[22] | BNT162b2 (1 dose) and ChAdOx1 nCoV-19 (1 dose) | *% female:* 51.7%<br>*Age:* mean 65 (vaccinated group).<br>*Country:* Scotland | *Type:* Cohort<br>*Comparison:* Unvaccinated controls. | *BNT162b2:* 711,839<br>*ChAdOx1 nCoV-19:* 620,154<br>*Unvaccinated:* 3,077,595 | 8-Dec-2020 to 22-Feb-2021 | From the first dose. Extracted case counts from day 14 post-first dose onwards.§ | Hospitalization |
| Baden et al (2021)[23] | mRNA-1273 (2 doses) | *% female:* 47.3%<br>*Age:* mean 51.4 (range: 18 to 95) | *Type:* RCT<br>*Comparison:* Saline placebo. | *Vaccinated:* 14,134<br>*Unvaccinated:* 14,073 | 27-July-2020 to 23-Oct-2020 | 14-days post-second dose. | Infection<br>Severe illness<br>Death |

| | | | | | | | |
|---|---|---|---|---|---|---|---|
| | | *Country:* United States. | | | | | |
| Emary et al (2021)[24]‡ | ChAdOx1 nCoV-19 (2 doses) | *% female:* 59.4%<br>*Age:* 18 to ≥70. Full range, means, or medians not reported.<br>*Country:* UK | *Type:* RCT<br>*Comparison:* MenACWY placebo. | *Vaccinated:* 4,244<br>*Unvaccinated:* 4,290 | 31-May-2020 to 30-Dec-2020 | 14-days post-second dose. | Infection<br>Hospitalization<br>Death |
| Madhi et al (2021)[25] | ChAdOx1 nCoV-19 (2 doses) | *% female:* 42.9%<br>*Age:* median 31 (IQR: 24 to 41)<br>*Country:* South Africa | *Type:* RCT<br>*Comparison:* Saline placebo | *Vaccinated:* 750<br>*Unvaccinated:* 717 | 24-June-2020 to 9-Nov-2020 | 14-days post-second dose. | Infection<br>Severe illness/hospitalization |
| Voysey et al (2021a)[26]‡ | ChAdOx1 nCoV-19 (2 doses) | *% female:* 60.5%<br>*Age:* 18 to ≥70. Full range, means, or medians not reported.<br>*Country:* UK, Brazil, South Africa. | *Type:* RCT<br>*Comparison:* MenACWY or saline placebo. | *Vaccinated:* 5,807<br>*Unvaccinated:* 5,829 | 23-April-2020 to 4-Nov-2020 | 14-days post-second dose. | Infection<br>Hospitalization<br>Death |
| Voysey et al (2021b)[27]‡ | ChAdOx1 nCoV-19 (2 doses) | *% female:* 56.4%<br>*Age:* 18 to ≥70. Full range, means, or medians not reported.<br>*Country:* UK, Brazil, South Africa. | *Type:* RCT<br>*Comparison:* MenACWY or saline placebo. | *Vaccinated:* 8,597<br>*Unvaccinated:* 8,581 | 23-April-2020 to 7-Dec-2020 | 14-days post-second dose. | Infection<br>Hospitalization<br>Death |
| Corchado-Garcia et al (2021)[28] | Ad26.COV2.S (1 dose) | *% female:* 53.8%<br>*Age:* 18 to ≥75. Full range, means, or medians not reported.<br>*Country:* United States | *Type:* Cohort<br>*Comparison:* Unvaccinated controls. | *Vaccinated:* 1,779<br>*Unvaccinated:* 17,744 | 27-Feb-2021 to 14-Apr-2021 | 15-days post-dose. | Infection<br>Hospitalization<br>Death |
| Sadoff et al (2021)[29] | Ad26.COV2.S (1 dose) | *% female:* 45%<br>*Age:* median 52 (range: 18 to 100)<br>*Country:* Argentina, Brazil, Chile, Colombia, Mexico, Peru, South Africa, United States. | *Type:* RCT<br>*Comparison:* Saline placebo. | *Vaccinated:* 19,514<br>*Unvaccinated:* 19,544 | 21-Sept-2020 to 22-Jan-2021 | 14-days post-dose. | Infection<br>Severe/critical illness<br>Death |
| Logunov et al (2021)[30] | Gam-COVID-Vac (2 doses) | *% female:* 38.8%<br>*Age:* mean 45.3<br>*Country:* Russia. | *Type:* RCT<br>*Comparison:* placebo made from the vaccine buffer composition. | *Vaccinated:* 14,964<br>*Unvaccinated:* 4,902 | 7-Sept-2020 to 24-Nov-2020 | 21-days post-first dose. | Infection<br>Moderate or severe illness<br>Death |
| Kaabi et al (2021)[31] | WIV04 (2 doses) and | *% female:* 15.6%<br>*Age:* mean 36.1 | *Type:* RCT | *WIV04:* 12,743<br>*HB02:* 12,726 | 16-July-2020 to 20-Dec-2020 | 14-days post-second dose. | Infection<br>Severe illness |

| | | | | | | | |
|---|---|---|---|---|---|---|---|
| | HB02 (2 doses) | *Country:* UAE, India, Bangladesh, China, Pakistan, Bahrain, Egypt, Philippines, Nepal, Syria, “others”. | *Comparison:* aluminium hydroxide (alum)-only placebo. | *Unvaccinated:* 12,737 | | | Death |
| Shinde et al (2021)[32] | NVX-CoV2373 (2 doses) | *% female:* 42.6%<br>*Age:* mean 32 (range: 18 to 84)<br>*Country:* South Africa | *Type:* RCT<br>*Comparison:* Saline placebo. | *Vaccinated:* 1,357<br>*Unvaccinated:* 1,327 | 17-Aug-2020 to 25-Nov-2020 | 7-days post-second dose. | Infection<br>Severe illness |

MenACWY=meningococcal group A, C, W, and Y conjugate vaccine; RCT=randomized controlled trial. Sample sizes are the totals for the infection data. If there was no infection data, the sample sizes are the totals for the severe illness/hospitalization or death data.

†Approximated unvaccinated sample size by taking the difference (*n*=1,895) between the number of people who received at least one vaccine (*n*=7,214) and the total sample size (*n*=9,109). Utilized this approximation because the authors reported totals as person-days. This approximation is an underestimate given that a portion of the unvaccinated person-days included the period before people were vaccinated (79% of the total sample would eventually become vaccinated). Thus, it is a conservative estimate since it means the estimated baseline infection risk is higher using this denominator (89/1,895=4.7%).

††Vaccinated sample was a group of healthcare workers (n=2,590) but the authors report 81% were vaccinated (n= 2,116). Cases were not reported separately for the vaccinated subgroup of healthcare workers. Therefore, we assumed all reported infections were from this vaccinated subgroup by using the number of vaccinated healthcare workers in the denominator.

‡Partially overlapping samples.

‡‡Approximated the vaccinated and unvaccinated sample sizes by extracting the total number of tests rather than cases. Vaccines were combined because the total number of tests was not reported separately by vaccine. Utilized this approximation because the authors reported totals as person-days. They also reported the number of people who received BNT162b2 (*n*=3,022), ChAdOx1 nCoV-19 (*n*=6,138), the total sample size (*n*=10,412), the never-vaccinated group (*n*=1,252), and the number of positive tests in the never-vaccinated group (*n*=328). However, because these were not unique tests, this denominator would yield a very high baseline infection risk (26.2%). Therefore, approximating the sample sizes using the overall number of tests yielded a less distorted baseline infection risk (723/15,392=4.7%) more consistent with the other studies.

§Authors report three different total number of events for the unvaccinated group: overall comparison (*n*=7,698), BNT162b2 comparison (*n*=6,671), and the ChAdOx1 nCov-19 comparison (*n*=7,252). Extracted the overall comparison since it was the higher number. Extracted event counts for the vaccine group from day 14 post-first dose onwards (day 14 to 42+ days). In the ChAdOx1 nCoV-19 group at days 35-41 and 42+, the counts are reported as ≤5. Therefore, we coded them as 5.

§§Authors reported total number of tests rather than cases.

∗Authors reported the number at risk and cumulative case counts at 0, 7, 14, 21, 28, 35, and 42 days post-first dose separately for SARS-CoV-2 infection, hospitalization, and death. Extracted the numbers at risk for vaccinated/unvaccinated groups at the start of day 28 until end of follow-up, which was 7 days post-second dose (patients received the second dose on day 21 post-first dose). These were the sizes of the vaccinated/unvaccinated groups for each of these outcomes. Extracted the counts for the new cases during this same period by taking the cumulative number of cases at day 42 minus the cumulative number of cases at day 28 for each group.

∗∗Authors reported the fully vaccinated population (*n*=4,714,932) and the Israel population aged 16 years and older (*n*=6,538,911). Approximated the unvaccinated population by taking the difference between these two numbers.

**Table S2.** Excluded studies identified in recent systematic reviews[1-3]

| Study | Vaccines | Reason for exclusion |
|---|---|---|
| Abu-Raddad et al (2021)[33] | BNT162b2 | Case-control study. |
| Andrejko et al (2021)[34] | BNT162b2 and mRNA-1273 | Case-control study. |
| Britton et al (2021)[35] | BNT162b2 | This was a small study of nursing home residents (*N*=463) at two facilities experiencing a SARS-CoV-2 outbreak. Thus, this study was more a study of transmission than efficacy against SARS-CoV-2 outcomes in the general population. Hence the very high baseline risk (44.8%). However, authors did not report the vaccination status of the index case. |
| Chodick et al (2021)[36] | BNT162b2 | No unvaccinated comparison. Compared infection rates from days 1 to 12 vs. days 13 to 24 post-first dose. |
| European Medicines Agency[37] | Ad26.COV2.S | This appears to be an earlier report of Sadoff et al (2021).[29] |
| Frenck et al (2021)[38] | BNT162b2 | Adolescent sample. |
| Gras-Valentí et al (2021)[39] | BNT162b2 | Case-control study. |
| Hall et al (2021)[40] | BNT162b2 | Authors report totals as person-days. Moreover, while they report the total size of never-vaccinated (*n*=2,683) and vaccinated (*n*=20,641) groups, the number of infections (*n*=977) in the unvaccinated cohort is not reported separately for these two groups. An unknown number of these infections were censored cases who would eventually be vaccinated, since a portion of the unvaccinated person-days included the period before people were vaccinated. This is suggested by the very high baseline infection risk if we used the number of never-vaccinated in the denominator (36.4%). Conversely, approximating the sample sizes using person-days (*n*=710,587) would yield a very low baseline risk (0.14%). Given the uncertainty and risk of distortion in the estimated baseline infection risk, we excluded this study. |
| Lopez Bernal et al (2021)[41] | BNT162b2 and ChAdOx1 nCoV-19 | Case-control study. |
| Martínez-Baz et al (2021)[42] | BNT162b2 mRNA-1273, and ChAdOx1 nCoV-19 | This study was of people who had close contact with a person with confirmed SARS-CoV-2 infection. 50.4% were household contacts. Thus, this study was more a study of transmission than efficacy against SARS-CoV-2 outcomes in the general population. Hence the very high baseline risk (35.7%). However, authors did not report the vaccination status of the index case. Notably, this appears to be an overlapping sample with the dedicated transmission study included in the meta-analysis of transmission risk.[43] |
| Mason et al (2021)[44] | BNT162b2 | Case-control study. |
| Pawlowski et al (2021)[45] | BNT162b2 and mRNA-1273 | Case-control study. |
| Pilishvili et al (2021)[46] | BNT162b2 and mRNA-1273 | Case-control study. |
| Tenforde et al (2021)[47] | BNT162b2 and mRNA-1273 | Case-control study. |
| Thompson et al (2021)[48] | BNT162b2 and mRNA-1273 | Authors report totals as person-days. Moreover, while they report the total size of never-vaccinated (*n*=989) and vaccinated (*n*=2,961) groups, the number of infections (*n*=161) in the unvaccinated cohort is not reported separately for these two groups. An unknown number of these infections |

| | |
|---|---|
| | were censored cases who would eventually be vaccinated, since a portion of the unvaccinated person-days included the period before people were vaccinated. This is suggested by the very high baseline infection risk if we used the number of never-vaccinated in the denominator (16.3%). Conversely, approximating the sample sizes using person-days (*n*=116,657) would yield a very low baseline risk (0.14%). Given the uncertainty and risk of distortion in the estimated baseline infection risk, we excluded this study. |

**Table S3.** Meta-analysis of SARS-CoV-2 vaccines on infection in the general population

| Vaccine | Sample size | Baseline (unvaccinated) infection risk† | Breakthrough (vaccinated) infection risk‡ | Infection RRR (95% CI) | Infection ARR (95% CI) | Infection NNV (95% CI) |
|---|---|---|---|---|---|---|
| ***Overall vaccine effect*** | *V:* 5,575,049<br>*UV:* 4,341,745 | 3.04%<br>(2.08%, 4.01%) | 0.57%<br>(0.27%, 0.87%) | 88%<br>(83%, 92%) | 2.59%<br>(1.75%, 3.43%) | 39<br>(29, 57) |
| **Heterogeneity** | --- | $I^2$ = 99.98%<br>Q ($df$ = 26) =<br>134,628.18<br>$p$ < 0.0001 | $I^2$ = 99.97%<br>Q ($df$ = 30) =<br>4,532.34<br>$p$ < 0.0001 | $I^2$ = 99.65%<br>Q ($df$ = 30) =<br>16,735.58<br>$p$ < 0.0001 | $I^2$ = 99.95%<br>Q ($df$ = 30) =<br>71,662.68<br>$p$ < 0.0001 | --- |
| **BNT162b2** | | | | | | |
| Amit et al (2021)[4] | *V:* 7,214<br>*UV:* 1,895 | 4.70% | 0.36% | 92%<br>(88%, 95%) | 4.34%<br>(3.37%, 5.30%) | 23<br>(19, 30) |
| Björk et al (2021)[5] | *V:* 26,587<br>*UV:* 779,154 | 0.53% | 0.16% | 70%<br>(59%, 78%) | 0.37%<br>(0.32%, 0.42%) | 269<br>(237, 312) |
| Dagan et al (2021)[6] | *V:* 108,529<br>*UV:* 107,209 | 0.30% | 0.05% | 83%<br>(78%, 87%) | 0.25%<br>(0.22%, 0.29%) | 396<br>(347, 461) |
| Fabiani et al (2021)[7] | *V:* 5,186<br>*UV:* 1,090 | 1.38% | 0.08% | 94%<br>(83%, 98%) | 1.30%<br>(0.60%, 1.99%) | 77<br>(50, 166) |
| Guijarro et al (2021)[8] | *V:* 2,116<br>*UV:* 170,513 | 2.69% | 0.28% | 89%<br>(77%, 95%) | 2.40%<br>(2.16%, 2.64%) | 42<br>(38, 46) |
| Haas et al (2021)[9] | *V:* 4,714,932<br>*UV:* 1,823,979 | 6.02% | 0.13% | 98%<br>(98%, 98%) | 5.89%<br>(5.86%, 5.93%) | 17<br>(17, 17) |
| Jones et al (2021)[10] | *V:* 1,989<br>*UV:* 3,252 | 0.80% | 0.20% | 75%<br>(28%, 91%) | 0.60%<br>(0.23%, 0.96%) | 167<br>(104, 427) |
| Khan & Mahmud (2021)[15] | *V:* 2,873<br>*UV:* 14,697 | 1.34% | 0.10% | 92%<br>(76%, 98%) | 1.24%<br>(1.02%, 1.46%) | 81<br>(69, 98) |
| Menni et al (2021)[19] | *V:* 67,293<br>*UV:* 464,356 | 10.84% | 3.66% | 66%<br>(65%, 68%) | 7.18%<br>(7.01%, 7.35%) | 14<br>(14, 14) |
| Monge et al (2021)[11] | *V:* 207,774<br>*UV:* 230,438 | 3.21% | 0.43% | 87%<br>(86%, 88%) | 2.78%<br>(2.70%, 2.86%) | 36<br>(35, 37) |
| Moustsen-Helms et al (2021)[12] | *V:* 114,406<br>*UV:* 255,673 | 2.41% | 0.03% | 99%<br>(98%, 99%) | 2.37%<br>(2.31%, 2.43%) | 42<br>(41, 43) |
| Polack et al (2020)[13] | *V:* 18,198<br>*UV:* 18,325 | 0.88% | 0.04% | 95%<br>(90%, 98%) | 0.84%<br>(0.70%, 0.98%) | 119<br>(102, 143) |
| Pritchard et al (2021)[20] | *V:* 57,646<br>*UV:* 329,419 | 3.25% | 0.12% | 96%<br>(95%, 97%) | 3.13%<br>(3.06%, 3.20%) | 32<br>(31, 33) |
| Tang et al (2021)[14] | *V:* 2,724<br>*UV:* 2,165 | 8.55% | 0.22% | 97%<br>(94%, 99%) | 8.32%<br>(7.13%, 9.52%) | 12<br>(11, 14) |
| Vasileiou et al (2021)[22] | *V:* 711,839<br>*UV:* 3,077,595 | *n.d.* | *n.d.* | *n.d.* | *n.d.* | *n.d.* |
| ***Vaccine effect*** | *V:* 5,337,467<br>*UV:* 4,202,165 | 3.34%<br>(1.68%, 5.00%) | 0.42%<br>(-0.07%, 0.91%) | 92%<br>(87%, 96%) | 2.91%<br>(1.56%, 4.25%) | 34<br>(24, 64) |
| **Heterogeneity** | --- | $I^2$ = 99.99%<br>Q ($df$ = 13) =<br>133,042.64<br>$p$ < 0.0001 | $I^2$ = 99.99%<br>Q ($df$ = 13) =<br>3,320.82<br>$p$ < 0.0001 | $I^2$ = 99.74%<br>Q ($df$ = 13) =<br>13,931.06<br>$p$ < 0.0001 | $I^2$ = 99.98%<br>Q ($df$ = 13) =<br>62,858.07<br>$p$ < 0.0001 | --- |
| **mRNA-1273** | | | | | | |
| Baden et al (2021)[23] | *V:* 14,134<br>*UV:* 14,073 | 1.31% | 0.08% | 94%<br>(89%, 97%) | 1.24%<br>(1.04%, 1.43%) | 81<br>(70, 96) |
| Khan & Mahmud (2021)[15] | *V:* 3,380<br>*UV:* 14,697 | 1.34% | 0.12% | 91%<br>(76%, 97%) | 1.22%<br>(1.00%, 1.44%) | 82<br>(69, 100) |
| ***Vaccine effect*** | *V:* 17,514<br>*UV:* 28,770 | 1.33%<br>(1.20%, 1.46%) | 0.08%<br>(0.04%, 0.13%) | 93%<br>(89%, 96%) | 1.23%<br>(1.09%, 1.38%) | 81<br>(73, 92) |

| | | | | | | |
|---|---|---|---|---|---|---|
| **Heterogeneity** | --- | $I^2$ = 0.00%<br>Q ($df$ = 1) =<br>0.0366<br>$p$ = 0.8482 | $I^2$ = 0.00%<br>Q ($df$ = 1) =<br>0.4056<br>$p$ = 0.5242 | $I^2$ = 0.00%<br>Q ($df$ = 1) =<br>0.4552<br>$p$ = 0.4999 | $I^2$ = 0.00%<br>Q ($df$ = 1) =<br>0.0097<br>$p$ = 0.9216 | --- |
| **ChAdOx1 nCoV-19** | | | | | | |
| Emary et al (2021)[24]‡ | *V:* 4,244<br>*UV:* 4,290 | 4.90% | 1.39% | 72%<br>(62%, 79%) | 3.50%<br>(2.77%, 4.24%) | 29<br>(24, 36) |
| Madhi et al (2021)[25] | *V:* 750<br>*UV:* 717 | 3.21% | 2.53% | 21%<br>(-44%, 57%) | 0.67%<br>(-1.04%, 2.39%) | 148<br>(42, -96) |
| Menni et al (2021)[19] | *V:* 36,329<br>*UV:* 464,356 | 10.84% | 1.76% | 84%<br>(82%, 85%) | 9.08%<br>(8.91%, 9.24%) | 11<br>(11, 11) |
| Pritchard et al (2021)[20] | *V:* 41,018<br>*UV:* 329,419 | 3.25% | 0.05% | 98%<br>(98%, 99%) | 3.20%<br>(3.14%, 3.27%) | 31<br>(31, 32) |
| Vasileiou et al (2021)[22] | *V:* 620,154<br>*UV:* 3,077,595 | *n.d.* | *n.d.* | *n.d.* | *n.d.* | *n.d.* |
| Voysey et al (2021a)[26]‡ | *V:* 5,807<br>*UV:* 5,829 | 1.73% | 0.52% | 70%<br>(55%, 80%) | 1.22%<br>(0.83%, 1.60%) | 82<br>(63, 120) |
| Voysey et al (2021b)[27]‡ | *V:* 8,597<br>*UV:* 8,581 | 2.89% | 0.98% | 66%<br>(57%, 74%) | 1.91%<br>(1.50%, 2.32%) | 52<br>(43, 67) |
| ***Vaccine effect*** | *V:* 96,745<br>*UV:* 813,192 | 4.48%<br>(1.83%, 7.12%) | 1.12%<br>(0.47%, 1.77%) | 80%<br>(43%, 93%) | 3.30%<br>(0.85%, 5.75%) | 30<br>(17, 118) |
| **Heterogeneity** | --- | $I^2$ = 99.94%<br>Q ($df$ = 5) =<br>19,688.69<br>$p$ < 0.0001 | $I^2$ = 99.06%<br>Q ($df$ = 5) =<br>755.22<br>$p$ < 0.0001 | $I^2$ = 99.00%<br>Q ($df$ = 5) =<br>194.67<br>$p$ < 0.0001 | $I^2$ = 99.85%<br>Q ($df$ = 5) =<br>4,653.06<br>$p$ < 0.0001 | --- |
| **Ad26.COV2.S** | | | | | | |
| Corchado-Garcia et al (2021)[28] | *V:* 1,779<br>*UV:* 17,744 | 0.72% | 0.17% | 77%<br>(27%, 93%) | 0.55%<br>(0.33%, 0.78%) | 181<br>(128, 308) |
| Sadoff et al (2021)[29] | *V:* 19,514<br>*UV:* 19,544 | 1.80% | 0.60% | 67%<br>(59%, 73%) | 1.20%<br>(0.98%, 1.41%) | 84<br>(71, 102) |
| ***Vaccine effect*** | *V:* 21,293<br>*UV:* 37,288 | 1.26%<br>(0.20%, 2.31%) | 0.39%<br>(-0.03%, 0.81%) | 67%<br>(59%, 73%) | 0.88%<br>(0.24%, 1.51%) | 114<br>(66, 408) |
| **Heterogeneity** | --- | $I^2$ = 98.87%<br>Q ($df$ = 1) =<br>88.41<br>$p$ < 0.0001 | $I^2$ = 93.26%<br>Q ($df$ = 1) =<br>14.84<br>$p$ < 0.0001 | $I^2$ = 0.00%<br>Q ($df$ = 1) =<br>0.3609<br>$p$ = 0.548 | $I^2$ = 93.83%<br>Q ($df$ = 1) =<br>16.20<br>$p$ < 0.0001 | --- |
| **Gam-COVID-Vac** | | | | | | |
| Logunov et al (2021)[30] | *V:* 14,964<br>*UV:* 4,902 | 1.26% | 0.11% | 92%<br>(85%, 95%) | 1.16%<br>(0.84%, 1.48%) | 86<br>(68, 119) |
| **WIV04** | | | | | | |
| Kaabi et al (2021)[31] | *V:* 12,743<br>*UV:* 12,737 | 0.75% | 0.20% | 73%<br>(58%, 82%) | 0.54%<br>(0.37%, 0.71%) | 185<br>(141, 268) |
| **HB02** | | | | | | |
| Kaabi et al (2021)[31] | *V:* 12,726<br>*UV:* 12,737 | 0.75% | 0.17% | 78%<br>(65%, 86%) | 0.58%<br>(0.42%, 0.75%) | 172<br>(134, 241) |
| **NVX-CoV2373** | | | | | | |
| Shinde et al (2021)[32] | *V:* 1,357<br>*UV:* 1,327 | 2.19% | 1.11% | 49%<br>(6%, 73%) | 1.08%<br>(0.12%, 2.04%) | 93<br>(49, 858) |
| **Mixed vaccines** | | | | | | |
| Glampson et al (2021)[17], BNT162b2 and ChAdOx1 nCoV-19 | *V:* 389,587<br>*UV:* 1,794,352 | *n.d.* | *n.d.* | *n.d.* | *n.d.* | *n.d.* |
| Lumley et al (2021)[18], BNT162b2 and ChAdOx1 nCoV-19 | *V:* 940<br>*UV:* 10,513 | 6.04% | 0.21% | 96%<br>(86%, 99%) | 5.83%<br>(5.29%, 6.37%) | 17<br>(16, 19) |

| | | | | | | |
|---|---|---|---|---|---|---|
| Shrotri et al (2021)[21], BNT162b2 and ChAdOx1 nCoV-19 | *V:* 15,289<br>*UV:* 15,392 | 4.70% | 2.41% | 49%<br>(42%, 55%) | 2.29%<br>(1.88%, 2.70%) | 44<br>(37, 53) |
| Swift et al (2021)[16], BNT162b2 and mRNA-1273 | *V:* 44,011<br>*UV:* 23,931 | 4.17% | 0.07% | 98%<br>(98%, 99%) | 4.10%<br>(3.84%, 4.35%) | 24<br>(23, 26) |
| ***Vaccine effect*** | *V:* 60,240<br>*UV:* 49,836 | 4.95%<br>(3.87%, 6.04%) | 0.89%<br>(-0.59%, 2.38%) | 93%<br>(42%, 99%) | 4.07%<br>(2.07%, 6.06%) | 25<br>(17, 48) |
| **Heterogeneity** | --- | $I^2 = 96.76\%$<br>Q ($df$ = 2) = 49.88<br>$p < 0.0001$ | $I^2 = 99.46\%$<br>Q ($df$ = 2) = 353.12<br>$p < 0.0001$ | $I^2 = 98.94\%$<br>Q ($df$ = 2) = 320.88<br>$p < 0.0001$ | $I^2 = 98.72\%$<br>Q ($df$ = 2) = 109.13<br>$p < 0.0001$ | --- |

'General population' means the denominators used the vaccinated vs. unvaccinated group sample sizes. V=vaccinated, UV=unvaccinated. Overall vaccine effect=meta-analysis of all effects across studies. Vaccine effect=meta-analysis of all effects for a given vaccine. RRR=relative risk reduction (1 – relative risk). ARR=absolute risk reduction. NNV=number needed to vaccinate. 95% CI=95% confidence interval. Brackets are the 95% confidence intervals (lower limit, upper limit). *Note:* displayed risks, RRRs, and ARRs are rounded to two decimal places but were calculated from unrounded numbers. Further, the ARR is the meta-analytic weighted average of the risk differences within studies, not the difference of the overall averages. Thus, the ARR is not simply the overall unvaccinated risk minus the overall vaccinated risk. *n.d.*=no data

†Baseline infection risk is defined as the SARS-CoV-2 infection risk in the unvaccinated population. The meta-analysis of the baseline infection risks across all studies excluded overlapping study populations because some studies compared two vaccines against the same unvaccinated population: Kaabi et al (2021),[31] Khan & Mahmud (2021),[15] Menni et al (2021),[19] and Pritchard et al (2021).[20] It also excluded studies with no infection data: Vasileiou et al (2021)[22] and Glampson et al (2021).[17] The meta-analysis of the baseline infection risks for each vaccine excluded only those studies with no infection data.

‡Breakthrough infection risk is the risk of SARS-CoV-2 infection post-vaccination. The meta-analysis of the overall effect and vaccine effects included all studies except those with no infection data.

**Table S4.** Meta-analysis of SARS-CoV-2 vaccines on severe illness/hospitalization in the general population

| Vaccine | Sample size | Baseline (unvaccinated) risk† | Breakthrough (vaccinated) risk‡ | Severe illness/hospitalization RRR (95% CI) | ARR (95% CI) | NNV (95% CI) |
|---|---|---|---|---|---|---|
| ***Overall vaccine effect*** | *V:* 6,287,289<br>*UV:* 5,258,797 | 0.16%<br>(0.09%, 0.23%) | 0.01%<br>(0.00%, 0.01%) | 89%<br>(82%, 93%) | 0.15%<br>(0.09%, 0.20%) | 676<br>(489, 1,094) |
| **Heterogeneity** | --- | $I^2 = 99.69\%$<br>Q ($df = 13$) = 5,868.93<br>$p < 0.0001$ | Cannot meaningfully calculate heterogeneity§ | $I^2 = 93.46\%$<br>Q ($df = 16$) = 184.34<br>$p < 0.0001$ | $I^2 = 99.51\%$<br>Q ($df = 16$) = 3,685.75<br>$p < 0.0001$ | --- |
| **BNT162b2** | | | | | | |
| Amit et al (2021)[4] | *n.d.* | *n.d.* | *n.d.* | *n.d.* | *n.d.* | *n.d.* |
| Björk et al (2021)[5] | *n.d.* | *n.d.* | *n.d.* | *n.d.* | *n.d.* | *n.d.* |
| Dagan et al (2021)[6] | *V:* 110,054<br>*UV:* 109,867 | 0.01% | 0.00% | 87%<br>(42%, 97%) | 0.01%<br>(0.00%, 0.02%) | 8,449<br>(5,211, 22,311) |
| Fabiani et al (2021)[7] | *n.d.* | *n.d.* | *n.d.* | *n.d.* | *n.d.* | *n.d.* |
| Guijarro et al (2021)[8] | *n.d.* | *n.d.* | *n.d.* | *n.d.* | *n.d.* | *n.d.* |
| Haas et al (2021)[9] | *V:* 4,714,932<br>*UV:* 1,823,979 | 0.30% | 0.01% | 96%<br>(95%, 96%) | 0.29%<br>(0.28%, 0.30%) | 344<br>(335, 354) |
| Jones et al (2021)[10] | *n.d.* | *n.d.* | *n.d.* | *n.d.* | *n.d.* | *n.d.* |
| Khan & Mahmud (2021)[15] | *V:* 2,873<br>*UV:* 14,697 | 0.32% | 0.03% | 89%<br>(21%, 98%) | 0.28%<br>(0.17%, 0.40%) | 351<br>(251, 585) |
| Menni et al (2021)[19] | *n.d.* | *n.d.* | *n.d.* | *n.d.* | *n.d.* | *n.d.* |
| Monge et al (2021)[11] | *n.d.* | *n.d.* | *n.d.* | *n.d.* | *n.d.* | *n.d.* |
| Moustsen-Helms et al (2021)[12] | *n.d.* | *n.d.* | *n.d.* | *n.d.* | *n.d.* | *n.d.* |
| Polack et al (2020)[13] | *V:* 21,669<br>*UV:* 21,686 | 0.04% | 0.00% | 89%<br>(12%, 99%) | 0.04%<br>(0.01%, 0.07%) | 2,711<br>(1,528, 12,035) |
| Pritchard et al (2021)[20] | *n.d.* | *n.d.* | *n.d.* | *n.d.* | *n.d.* | *n.d.* |
| Tang et al (2021)[14] | *n.d.* | *n.d.* | *n.d.* | *n.d.* | *n.d.* | *n.d.* |
| Vasileiou et al (2021)[22] | *V:* 711,839<br>*UV:* 3,077,595 | 0.25% | 0.03% | 90%<br>(88%, 91%) | 0.22%<br>(0.22%, 0.23%) | 444<br>(432, 458) |
| ***Vaccine effect*** | *V:* 5,561,367<br>*UV:* 5,047,824 | 0.18%<br>(0.05%, 0.31%) | 0.01%<br>(0.01%, 0.02%) | 92%<br>(86%, 96%) | 0.17%<br>(0.05%, 0.29%) | 601<br>(350, 2,125) |
| **Heterogeneity** | --- | $I^2 = 99.89\%$<br>Q ($df = 4$) = 3,840.83<br>$p < 0.0001$ | $I^2 = 93.86\%$<br>Q ($df = 4$) = 116.22<br>$p < 0.0001$ | $I^2 = 94.41\%$<br>Q ($df = 4$) = 104.75<br>$p < 0.0001$ | $I^2 = 99.85\%$<br>Q ($df = 4$) = 2,984.53<br>$p < 0.0001$ | --- |
| **mRNA-1273** | | | | | | |
| Baden et al (2021)[23] | *V:* 14,134<br>*UV:* 14,073 | 0.21% | 0.00% | 98%<br>(73%, 100%) | 0.21%<br>(0.14%, 0.29%) | 469<br>(346, 730) |
| Khan & Mahmud (2021)[15] | *V:* 3,380<br>*UV:* 14,697 | 0.32% | 0.06% | 81%<br>(24%, 96%) | 0.26%<br>(0.14%, 0.38%) | 384<br>(261, 725) |
| ***Vaccine effect*** | *V:* 17,514<br>*UV:* 28,770 | 0.26%<br>(0.16%, 0.37%) | 0.01%<br>(-0.04%, 0.07%) | 92%<br>(25%, 99%) | 0.23%<br>(0.16%, 0.29%) | 442<br>(344, 619) |
| **Heterogeneity** | --- | $I^2 = 67.62\%$<br>Q ($df = 1$) = 3.0886<br>$p = 0.0788$ | Cannot meaningfully calculate heterogeneity§ | $I^2 = 56.68\%$<br>Q ($df = 1$) = 2.3086<br>$p = 0.1287$ | $I^2 = 0.00\%$<br>Q ($df = 1$) = 0.4146<br>$p = 0.5196$ | --- |
| **ChAdOx1 nCoV-19** | | | | | | |
| Emary et al (2021)[24]‡ | *i.d.* | *i.d.* | *i.d.* | *i.d.* | *i.d.* | *i.d.* |
| Madhi et al (2021)[25] | *i.d.* | *i.d.* | *i.d.* | *i.d.* | *i.d.* | *i.d.* |
| Menni et al (2021)[19] | *n.d.* | *n.d.* | *n.d.* | *n.d.* | *n.d.* | *n.d.* |
| Pritchard et al (2021)[20] | *n.d.* | *n.d.* | *n.d.* | *n.d.* | *n.d.* | *n.d.* |

| | | | | | | |
|---|---|---|---|---|---|---|
| Vasileiou et al (2021)[22] | *V:* 620,154<br>*UV:* 3,077,595 | 0.25% | 0.02% | 91%<br>(89%, 92%) | 0.23%<br>(0.22%, 0.23%) | 439<br>(426, 452) |
| Voysey et al (2021a)[26]‡ | *V:* 12,021<br>*UV:* 11,724 | 0.09% | 0.00% | 95%<br>(21%, 100%) | 0.09%<br>(0.03%, 0.14%) | 1,172<br>(724, 3,081) |
| Voysey et al (2021b)[27]‡ | *V:* 11,794<br>*UV:* 11,776 | 0.13% | 0.00% | 97%<br>(46%, 100%) | 0.13%<br>(0.06%, 0.19%) | 785<br>(521, 1,588) |
| ***Vaccine effect*** | *V:* 643,969<br>*UV:* 3,101,095 | 0.16%<br>(0.06%, 0.26%) | 0.01%<br>(-0.01%, 0.02%) | 91%<br>(90%, 93%) | 0.15%<br>(0.06%, 0.24%) | 664<br>(420, 1,582) |
| **Heterogeneity** | --- | $I^2$ = 94.36%<br>Q ($df$ = 2) =<br>50.40<br>$p$ < 0.0001 | Cannot meaningfully calculate heterogeneity§ | $I^2$ = 0.00%<br>Q ($df$ = 2) =<br>0.6970<br>$p$ = 0.7058 | $I^2$ = 92.42%<br>Q ($df$ = 2) =<br>36.36<br>$p$ < 0.0001 | --- |
| **Ad26.COV2.S** | | | | | | |
| Corchado-Garcia et al (2021)[28] | *V:* 2,195<br>*UV:* 124,377 | 0.02% | 0.05% | -198%<br>(-2,127%, 60%) | -0.03%<br>(-0.12%, 0.06%) | -3,302<br>(1,688,<br>-835) |
| Sadoff et al (2021)[29] | *V:* 19,514<br>*UV:* 19,544 | 0.31% | 0.07% | 77%<br>(58%, 87%) | 0.24%<br>(0.15%, 0.32%) | 425<br>(311, 671) |
| ***Vaccine effect*** | *V:* 21,709<br>*UV:* 143,921 | 0.16%<br>(-0.13%, 0.44%) | 0.07%<br>(0.03%, 0.10%) | 31%<br>(-715%, 94%) | 0.10%<br>(-0.16%, 0.36%) | 973<br>(275, -635) |
| **Heterogeneity** | --- | $I^2$ = 98.15%<br>Q ($df$ = 1) =<br>53.92<br>$p$ < 0.0001 | $I^2$ = 0.00%<br>Q ($df$ = 1) =<br>0.2808<br>$p$ = 0.5962 | $I^2$ = 82.42%<br>Q ($df$ = 1) =<br>5.6873<br>$p$ = 0.0171 | $I^2$ = 94.30%<br>Q ($df$ = 1) =<br>17.54<br>$p$ < 0.0001 | --- |
| **Gam-COVID-Vac** | | | | | | |
| Logunov et al (2021)[30] | *V:* 14,964<br>*UV:* 4,902 | 0.41% | 0.00% | 99%<br>(87%, 100%) | 0.41%<br>(0.23%, 0.59%) | 245<br>(171, 436) |
| **WIV04** | | | | | | |
| Kaabi et al (2021)[31] | *V:* 12,743<br>*UV:* 12,737 | 0.02% | 0.00% | 80%<br>(-316%, 99%) | 0.02%<br>(-0.01%, 0.04%) | 6,368<br>(2,669,<br>-16,507) |
| **HB02** | | | | | | |
| Kaabi et al (2021)[31] | *V:* 12,726<br>*UV:* 12,737 | 0.02% | 0.00% | 80%<br>(-317%, 99%) | 0.02%<br>(-0.01%, 0.04%) | 6,368<br>(2,669,<br>-16,507) |
| **NVX-CoV2373** | | | | | | |
| Shinde et al (2021)[32] | *V:* 1,357<br>*UV:* 1,327 | 0.08% | 0.00% | 67%<br>(-699%, 99%) | 0.08%<br>(-0.07%, 0.22%) | 1,327<br>(448, -1,383) |
| **Mixed vaccines** | | | | | | |
| Glampson et al (2021)[17], BNT162b2 and ChAdOx1 nCoV-19 | *n.d.* | *n.d.* | *n.d.* | *n.d.* | *n.d.* | *n.d.* |
| Lumley et al (2021)[18], BNT162b2 and ChAdOx1 nCoV-19 | *V:* 940<br>*UV:* 10,513 | 0.15% | 0.00% | 66%<br>(-464%, 98%) | 0.15%<br>(0.08%, 0.23%) | 657<br>(441, 1287) |
| Shrotri et al (2021)[21], BNT162b2 and ChAdOx1 nCoV-19 | *n.d.* | *n.d.* | *n.d.* | *n.d.* | *n.d.* | *n.d.* |
| Swift et al (2021)[16], BNT162b2 and mRNA-1273 | *n.d.* | *n.d.* | *n.d.* | *n.d.* | *n.d.* | *n.d.* |
| ***Vaccine effect*** | Insufficient data to meta-analyze | | | Insufficient data to meta-analyze | | |

'General population' means the denominators used the vaccinated vs. unvaccinated group sample sizes. V=vaccinated, UV=unvaccinated. Overall vaccine effect=meta-analysis of all effects across studies. Vaccine effect=meta-analysis of all effects for a given vaccine. RRR=relative risk reduction (1 – relative risk). ARR=absolute risk reduction. NNV=number needed to vaccinate.

95% CI=95% confidence interval. Brackets are the 95% confidence intervals (lower limit, upper limit). *Note:* displayed risks, RRRs, and ARRs are rounded to two decimal places but were calculated from unrounded numbers. Further, the ARR is the meta-analytic weighted average of the risk differences within studies, not the difference of the overall averages. Thus, the ARR is not simply the overall unvaccinated risk minus the overall vaccinated risk. *n.d.*=no data. *i.d.*=insufficient data (i.e., there were no cases in the vaccinated *and* unvaccinated groups).

†Baseline risk is defined as the risk of serious illness/hospitalization from SARS-CoV-2 in the unvaccinated population. The meta-analysis of the baseline risks across all studies excluded overlapping study populations because some studies compared two vaccines against the same unvaccinated population: Kaabi et al (2021),[31] Khan & Mahmud (2021),[15] Menni et al (2021),[19] and Pritchard et al (2021).[20] It also excluded studies with no or insufficient data for the outcome. The meta-analysis of the baseline risks for each vaccine excluded only those studies with no or insufficient data for the outcome.

‡Breakthrough risk is the risk of serious illness/hospitalization from SARS-CoV-2 post-vaccination. The meta-analysis of the overall effect and vaccine effects included all studies except those with no or insufficient data for the outcome.

§Cannot meaningfully calculate heterogeneity metrics because there are samples with values of 0.00% leading to sample variances of 0.00.

**Table S5.** Meta-analysis of SARS-CoV-2 vaccines on death in the general population

| Vaccine | Sample size | Baseline (unvaccinated) risk† | Breakthrough (vaccinated) risk‡ | Death RRR (95% CI) | Death ARR (95% CI) | Death NNV (95% CI) |
|---|---|---|---|---|---|---|
| ***Overall vaccine effect*** | Insufficient data to meta-analyze | | | Insufficient data to meta-analyze | | |
| **BNT162b2** | | | | | | |
| Amit et al (2021)[4] | *n.d.* | *n.d.* | *n.d.* | *n.d.* | *n.d.* | *n.d.* |
| Björk et al (2021)[5] | *V:* 26,587<br>*UV:* 779,154 | 0.00% | 0.00% | 60%<br>(-554%, 98%) | 0.00%<br>(0.00%, 0.01%) | 21,643<br>(16,314, 32,143) |
| Dagan et al (2021)[6] | *V:* 110,101<br>*UV:* 110,008 | 0.00% | 0.00% | 60%<br>(-106%, 92%) | 0.00%<br>(0.00%, 0.01%) | 36,649<br>(13,439, -50,404) |
| Fabiani et al (2021)[7] | *n.d.* | *n.d.* | *n.d.* | *n.d.* | *n.d.* | *n.d.* |
| Guijarro et al (2021)[8] | *n.d.* | *n.d.* | *n.d.* | *n.d.* | *n.d.* | *n.d.* |
| Haas et al (2021)[9] | *V:* 4,714,932<br>*UV:* 1,823,979 | 0.04% | 0.00% | 93%<br>(91%, 94%) | 0.04%<br>(0.03%, 0.04%) | 2,757<br>(2,552, 2,998) |
| Jones et al (2021)[10] | *n.d.* | *n.d.* | *n.d.* | *n.d.* | *n.d.* | *n.d.* |
| Khan & Mahmud (2021)[15] | *n.d.* | *n.d.* | *n.d.* | *n.d.* | *n.d.* | *n.d.* |
| Menni et al (2021)[19] | *n.d.* | *n.d.* | *n.d.* | *n.d.* | *n.d.* | *n.d.* |
| Monge et al (2021)[11] | *n.d.* | *n.d.* | *n.d.* | *n.d.* | *n.d.* | *n.d.* |
| Moustsen-Helms et al (2021)[12] | *n.d.* | *n.d.* | *n.d.* | *n.d.* | *n.d.* | *n.d.* |
| Polack et al (2020)[13] | *i.d.* | *i.d.* | *i.d.* | *i.d.* | *i.d.* | *i.d.* |
| Pritchard et al (2021)[20] | *n.d.* | *n.d.* | *n.d.* | *n.d.* | *n.d.* | *n.d.* |
| Tang et al (2021)[14] | *n.d.* | *n.d.* | *n.d.* | *n.d.* | *n.d.* | *n.d.* |
| Vasileiou et al (2021)[22] | *n.d.* | *n.d.* | *n.d.* | *n.d.* | *n.d.* | *n.d.* |
| ***Vaccine effect*** | Insufficient data to meta-analyze | | | Insufficient data to meta-analyze | | |
| **mRNA-1273** | | | | | | |
| Baden et al (2021)[23] | *V:* 14,134<br>*UV:* 14,073 | 0.01% | 0.00% | 67%<br>(-715%, 99%) | 0.01%<br>(-0.01%, 0.02%) | 14,073<br>(4,755, -14,661) |
| Khan & Mahmud (2021)[15] | *n.d.* | *n.d.* | *n.d.* | *n.d.* | *n.d.* | *n.d.* |
| ***Vaccine effect*** | Insufficient data to meta-analyze | | | Insufficient data to meta-analyze | | |
| **ChAdOx1 nCoV-19** | | | | | | |
| Emary et al (2021)[24]‡ | *i.d.* | *i.d.* | *i.d.* | *i.d.* | *i.d.* | *i.d.* |
| Madhi et al (2021)[25] | *n.d.* | *n.d.* | *n.d.* | *n.d.* | *n.d.* | *n.d.* |
| Menni et al (2021)[19] | *n.d.* | *n.d.* | *n.d.* | *n.d.* | *n.d.* | *n.d.* |
| Pritchard et al (2021)[20] | *n.d.* | *n.d.* | *n.d.* | *n.d.* | *n.d.* | *n.d.* |
| Vasileiou et al (2021)[22] | *n.d.* | *n.d.* | *n.d.* | *n.d.* | *n.d.* | *n.d.* |
| Voysey et al (2021a)[26]‡ | *V:* 12,021<br>*UV:* 11,724 | 0.01% | 0.00% | 67%<br>(-698%, 99%) | 0.01%<br>(-0.01%, 0.03%) | 11,724<br>(3,961, -12,214) |
| Voysey et al (2021b)[27]‡ | *V:* 11,794<br>*UV:* 11,776 | 0.01% | 0.00% | 67%<br>(-717%, 99%) | 0.01%<br>(-0.01%, 0.03%) | 11776<br>(3,979, -12,268) |
| ***Vaccine effect*** | Insufficient data to meta-analyze | | | Insufficient data to meta-analyze | | |
| **Ad26.COV2.S** | | | | | | |
| Corchado-Garcia et al (2021)[28] | *V:* 2,195<br>*UV:* 124,377 | 0.00% | 0.00% | -1,788%<br>(-46,231%, 23%) | 0.00%<br>(0.00%, 0.00%) | 124,377<br>(42,020, -129,565) |
| Sadoff et al (2021)[29] | *V:* 19,514<br>*UV:* 19,544 | 0.03% | 0.00% | 91%<br>(-65%, 99%) | 0.03%<br>(0.00%, 0.05%) | 3909<br>(2,083, 31,627) |
| ***Vaccine effect*** | Insufficient data to meta-analyze | | | Insufficient data to meta-analyze | | |
| **Gam-COVID-Vac** | | | | | | |
| Logunov et al (2021)[30] | *i.d.* | *i.d.* | *i.d.* | *i.d.* | *i.d.* | *i.d.* |
| **WIV04** | | | | | | |

| | | | | | | |
|---|---|---|---|---|---|---|
| Kaabi et al (2021)[31] | *i.d.* | *i.d.* | *i.d.* | *i.d.* | *i.d.* | *i.d.* |
| **HB02** | | | | | | |
| Kaabi et al (2021)[31] | *i.d.* | *i.d.* | *i.d.* | *i.d.* | *i.d.* | *i.d.* |
| **NVX-CoV2373** | | | | | | |
| Shinde et al (2021)[32] | *n.d.* | *n.d.* | *n.d.* | *n.d.* | *n.d.* | *n.d.* |
| **Mixed vaccines** | | | | | | |
| Glampson et al (2021)[17], BNT162b2 and ChAdOx1 nCoV-19 | *V:* 389,587<br>*UV:* 1,794,352 | 0.00% | 0.00% | -22%<br>(-105%, 27%) | 0.00%<br>(0.00%, 0.00%) | -120,394<br>(67,293, -31,774) |
| Lumley et al (2021)[18], BNT162b2 and ChAdOx1 nCoV-19 | *n.d.* | *n.d.* | *n.d.* | *n.d.* | *n.d.* | *n.d.* |
| Shrotri et al (2021)[21], BNT162b2 and ChAdOx1 nCoV-19 | *n.d.* | *n.d.* | *n.d.* | *n.d.* | *n.d.* | *n.d.* |
| Swift et al (2021)[16], BNT162b2 and mRNA-1273 | *n.d.* | *n.d.* | *n.d.* | *n.d.* | *n.d.* | *n.d.* |
| ***Vaccine effect*** | Insufficient data to meta-analyze | | | Insufficient data to meta-analyze | | |

‘General population’ means the denominators used the vaccinated vs. unvaccinated group sample sizes. V=vaccinated, UV=unvaccinated. Overall vaccine effect=meta-analysis of all effects across studies. Vaccine effect=meta-analysis of all effects for a given vaccine. RRR=relative risk reduction (1 – relative risk). ARR=absolute risk reduction. NNV=number needed to vaccinate. 95% CI=95% confidence interval. Brackets are the 95% confidence intervals (lower limit, upper limit). *Note:* displayed risks, RRRs, and ARRs are rounded to two decimal places but were calculated from unrounded numbers. *n.d.*=no data. *i.d.*=insufficient data (i.e., there were no cases in the vaccinated *and* unvaccinated groups).

†Baseline risk is defined as the risk of death from SARS-CoV-2 in the unvaccinated population.

‡Breakthrough risk is the risk of death from SARS-CoV-2 post-vaccination.

**Table S6.** Meta-analysis of SARS-CoV-2 vaccines on severe illness/hospitalization among infected persons

| Vaccine | Sample size | Baseline (unvaccinated) risk† | Breakthrough (vaccinated) risk‡ | Severe illness/hospitalization RRR (95% CI) | ARR (95% CI) | NNV (95% CI) |
|---|---|---|---|---|---|---|
| ***Overall vaccine effect*** | *V:* 6,669⁎<br>*UV:* 111,761⁎ | 11.02%⁎<br>(6.31%, 15.72%) | 2.50%⁎<br>(0.06%, 4.94%) | 7%⁎<br>(-55%, 44%) | 5.64%⁎<br>(0.69%, 10.59%) | 18⁎<br>(9, 146) |
| **Heterogeneity** | --- | $I^2$ = 96.88%<br>Q ($df$ = 11) = 129.40<br>$p < 0.0001$ | Cannot meaningfully calculate heterogeneity§ | $I^2$ = 51.83%<br>Q ($df$ = 13) = 30.68<br>$p = 0.0038$ | $I^2$ = 94.29%<br>Q ($df$ = 13) = 190.11<br>$p < 0.0001$ | --- |
| **BNT162b2** | | | | | | |
| Amit et al (2021)[4] | *n.d.* | *n.d.* | *n.d.* | *n.d.* | *n.d.* | *n.d.* |
| Björk et al (2021)[5] | *n.d.* | *n.d.* | *n.d.* | *n.d.* | *n.d.* | *n.d.* |
| Dagan et al (2021)[6] | *V:* 55<br>*UV:* 325 | 4.62% | 3.64% | 21%<br>(-235%, 81%) | 0.98%<br>(-4.47%, 6.43%) | 102<br>(16, -22) |
| Fabiani et al (2021)[7] | *n.d.* | *n.d.* | *n.d.* | *n.d.* | *n.d.* | *n.d.* |
| Guijarro et al (2021)[8] | *n.d.* | *n.d.* | *n.d.* | *n.d.* | *n.d.* | *n.d.* |
| Haas et al (2021)[9] | *V:* 6,266<br>*UV:* 109,876 | 5.03% | 9.51% | -89%<br>(-105%, -74%) | -4.48%<br>(-5.22%, -3.74%) | -22<br>(-27, -19) |
| Jones et al (2021)[10] | *n.d.* | *n.d.* | *n.d.* | *n.d.* | *n.d.* | *n.d.* |
| Khan & Mahmud (2021)[15] | *V:* 3<br>*UV:* 197 | 23.86% | 33.33% | -40%<br>(-606%, 72%) | -9.48%<br>(-63.15%, 44.20%) | -11<br>(2, -2) |
| Menni et al (2021)[19] | *n.d.* | *n.d.* | *n.d.* | *n.d.* | *n.d.* | *n.d.* |
| Monge et al (2021)[11] | *n.d.* | *n.d.* | *n.d.* | *n.d.* | *n.d.* | *n.d.* |
| Moustsen-Helms et al (2021)[12] | *n.d.* | *n.d.* | *n.d.* | *n.d.* | *n.d.* | *n.d.* |
| Polack et al (2020)[13] | *V:* 8<br>*UV:* 162 | 5.56% | 12.50% | -125%<br>(-1,466%, 68%) | -6.94%<br>(-30.13%, 16.24%) | -14<br>(6, -3) |
| Pritchard et al (2021)[20] | *n.d.* | *n.d.* | *n.d.* | *n.d.* | *n.d.* | *n.d.* |
| Tang et al (2021)[14] | *n.d.* | *n.d.* | *n.d.* | *n.d.* | *n.d.* | *n.d.* |
| Vasileiou et al (2021)[22] | *n.d.* | *n.d.* | *n.d.* | *n.d.* | *n.d.* | *n.d.* |
| ***Vaccine effect*** | *V:* 6,332<br>*UV:* 110,560 | 9.44%<br>(0.65%, 18.23%) | 7.67%<br>(2.65%, 12.68%) | -89%<br>(-104%, -74%) | -2.79%<br>(-7.36%, 1.78%) | -36<br>(56, -14) |
| **Heterogeneity** | --- | $I^2$ = 98.34%<br>Q ($df$ = 3) = 38.6417<br>$p < 0.0001$ | $I^2$ = 54.89%<br>Q ($df$ = 3) = 6.1482<br>$p = 0.1046$ | $I^2$ = 0.00%<br>Q ($df$ = 3) = 1.565<br>$p = 0.6673$ | $I^2$ = 43.64%<br>Q ($df$ = 3) = 3.8725<br>$p = 0.2756$ | --- |
| **mRNA-1273** | | | | | | |
| Baden et al (2021)[23] | *V:* 11<br>*UV:* 185 | 16.22% | 0.00% | 75%<br>(-291%, 98%) | 16.22%<br>(10.90%, 21.53%) | 6<br>(5, 9) |
| Khan & Mahmud (2021)[15] | *V:* 4<br>*UV:* 197 | 23.86% | 50.00% | -110%<br>(-476%, 24%) | -26.14%<br>(-75.50%, 23.22%) | -4<br>(4, -1) |
| ***Vaccine effect*** | *V:* 15<br>*UV:* 382 | 19.91%<br>(12.43%, 27.40%) | 18.75%<br>(-28.69%, 66.19%) | -9%<br>(-637%, 84%) | 2.44%<br>(-36.46%, 41.33%) | 41<br>(2, -3) |
| **Heterogeneity** | --- | $I^2$ = 71.63%<br>Q ($df$ = 1) = 3.5251<br>$p = 0.0604$ | Cannot meaningfully calculate heterogeneity§ | $I^2$ = 50.35%<br>Q ($df$ = 1) = 2.0142<br>$p = 0.1558$ | $I^2$ = 64.24%<br>Q ($df$ = 1) = 2.7966<br>$p = 0.0945$ | --- |
| **ChAdOx1 nCoV-19** | | | | | | |

| | | | | | | |
|---|---|---|---|---|---|---|
| Emary et al (2021)[24]‡ | *i.d.* | *i.d.* | *i.d.* | *i.d.* | *i.d.* | *i.d.* |
| Madhi et al (2021)[25] | *i.d.* | *i.d.* | *i.d.* | *i.d.* | *i.d.* | *i.d.* |
| Menni et al (2021)[19] | *n.d.* | *n.d.* | *n.d.* | *n.d.* | *n.d.* | *n.d.* |
| Pritchard et al (2021)[20] | *n.d.* | *n.d.* | *n.d.* | *n.d.* | *n.d.* | *n.d.* |
| Vasileiou et al (2021)[22] | *n.d.* | *n.d.* | *n.d.* | *n.d.* | *n.d.* | *n.d.* |
| Voysey et al (2021a)[26]‡ | *V:* 30<br>*UV:* 101 | 9.90% | 0.00% | 84%<br>(-160%, 99%) | 9.90%<br>(4.08%, 15.73%) | 10<br>(6, 25) |
| Voysey et al (2021b)[27]‡ | *V:* 84<br>*UV:* 248 | 6.05% | 0.00% | 91%<br>(-56%, 99%) | 6.05%<br>(3.08%, 9.02%) | 17<br>(11, 32) |
| ***Vaccine effect*** | *V:* 114<br>*UV:* 349 | 7.13%<br>(3.74%, 10.51%) | 0.00%<br>(0.00%, 0.00%) | 88%<br>(11%, 98%) | 7.13%<br>(3.74%, 10.51%) | 14<br>(10, 27) |
| **Heterogeneity** | --- | $I^2$ = 25.06%<br>Q ($df$ = 1) =<br>1.3343<br>$p$ = 0.248 | Cannot meaningfully calculate heterogeneity§ | $I^2$ = 0.00%<br>Q ($df$ = 1) =<br>0.0623<br>$p$ = 0.8028 | $I^2$ = 25.06%<br>Q ($df$ = 1) =<br>1.3343<br>$p$ = 0.248 | --- |
| **Ad26.COV2.S** | | | | | | |
| Corchado-Garcia et al (2021)[28] | *V:* 13<br>*UV:* 130 | 14.62% | 7.69% | 47%<br>(-262%, 92%) | 6.92%<br>(-8.78%, 22.63%) | 14<br>(4, -11) |
| Sadoff et al (2021)[29] | *V:* 117<br>*UV:* 351 | 17.09% | 11.97% | 30%<br>(-20%, 59%) | 5.13%<br>(-1.95%, 12.21%) | 20<br>(8, -51) |
| ***Vaccine effect*** | *V:* 130<br>*UV:* 481 | 16.36%<br>(13.06%, 19.66%) | 11.36%<br>(5.91%, 16.81%) | 31%<br>(-16%, 59%) | 5.43%<br>(-1.02%, 11.88%) | 18<br>(8, -98) |
| **Heterogeneity** | --- | $I^2$ = 0.00%<br>Q ($df$ = 1) =<br>0.4505<br>$p$ = 0.5021 | $I^2$ = 0.00%<br>Q ($df$ = 1) =<br>0.287<br>$p$ = 0.5921 | $I^2$ = 0.00%<br>Q ($df$ = 1) =<br>0.0778<br>$p$ = 0.7802 | $I^2$ = 0.00%<br>Q ($df$ = 1) =<br>0.0417<br>$p$ = 0.8382 | --- |
| **Gam-COVID-Vac** | | | | | | |
| Logunov et al (2021)[30] | *V:* 16<br>*UV:* 62 | 32.26% | 0.00% | 91%<br>-42%, 99%) | 32.26%<br>(20.62%, 43.89%) | 3<br>(2, 5) |
| **WIV04** | | | | | | |
| Kaabi et al (2021)[31] | *V:* 26<br>*UV:* 95 | 2.11% | 0.00% | 29%<br>(-1337%, 96%) | 2.11%<br>(-0.78%, 4.99%) | 48<br>(20, -128) |
| **HB02** | | | | | | |
| Kaabi et al (2021)[31] | *V:* 21<br>*UV:* 95 | 2.11% | 0.00% | 13%<br>(-1654%, 96%) | 2.11%<br>(-0.78%, 4.99%) | 48<br>(20, -128) |
| **NVX-CoV2373** | | | | | | |
| Shinde et al (2021)[32] | *n.d.* | *n.d.* | *n.d.* | *n.d.* | *n.d.* | *n.d.* |
| **Mixed vaccines** | | | | | | |
| Glampson et al (2021)[17], BNT162b2 and ChAdOx1 nCoV-19 | *n.d.* | *n.d.* | *n.d.* | *n.d.* | *n.d.* | *n.d.* |
| Lumley et al (2021)[18], BNT162b2 and ChAdOx1 nCoV-19 | *V:* 2<br>*UV:* 635 | 2.52% | 0.00% | -542%<br>(-8,334%, 51%) | 2.52%<br>(1.30%, 3.74%) | 40<br>(27, 77) |
| Shrotri et al (2021)[21], BNT162b2 and ChAdOx1 nCoV-19 | *n.d.* | *n.d.* | *n.d.* | *n.d.* | *n.d.* | *n.d.* |
| Swift et al (2021)[16], BNT162b2 and mRNA-1273 | *n.d.* | *n.d.* | *n.d.* | *n.d.* | *n.d.* | *n.d.* |
| ***Vaccine effect*** | Insufficient data to meta-analyze | | | Insufficient data to meta-analyze | | |

‘Among infected persons’ means the denominators used the number of infections in the vaccinated vs. unvaccinated groups. V=vaccinated, UV=unvaccinated. Overall vaccine effect=meta-analysis of all effects across studies. Vaccine effect=meta-analysis of all effects for a given vaccine. RRR=relative risk reduction (1 – relative risk). ARR=absolute risk reduction. NNV=number needed to vaccinate. 95% CI=95% confidence interval. Brackets are the 95% confidence intervals (lower limit, upper limit). *Note:* displayed risks, RRRs, and ARRs are rounded to two decimal places but were calculated from unrounded numbers. Further, the ARR is the meta-analytic weighted average of the risk differences within studies, not the difference of the overall averages. Thus, the ARR is not simply the overall unvaccinated risk minus the overall vaccinated risk. *n.d.*=no data. *i.d.*=insufficient data (i.e., there were no cases in the vaccinated *and* unvaccinated groups).
†Baseline risk is defined as the risk of serious illness/hospitalization from SARS-CoV-2 in the unvaccinated population. The meta-analysis of the baseline risks across all studies excluded overlapping study populations because some studies compared two vaccines against the same unvaccinated population: Kaabi et al (2021),[31] Khan & Mahmud (2021),[15] Menni et al (2021),[19] and Pritchard et al (2021).[20] It also excluded studies with no or insufficient data for the outcome. The meta-analysis of the baseline risks for each vaccine excluded only those studies with no or insufficient data for the outcome.
‡Breakthrough risk is the risk of serious illness/hospitalization from SARS-CoV-2 post-vaccination. The meta-analysis of the overall effect and vaccine effects included all studies except those with no or insufficient data for the outcome.
⁎Removed Lumley et al (2021)[18] since this was a significant outlier (RRR=-542) and had a small vaccinated group (*n*=2).
§Cannot meaningfully calculate heterogeneity metrics because there are samples with values of 0.00% leading to sample variances of 0.00.

**Table S7.** Meta-analysis of SARS-CoV-2 vaccines on death among infected persons

| Vaccine | Sample size | Baseline (unvaccinated) risk† | Breakthrough (vaccinated) risk‡ | Death RRR (95% CI) | Death ARR (95% CI) | Death NNV (95% CI) |
|---|---|---|---|---|---|---|
| ***Overall vaccine effect*** | Insufficient data to meta-analyze | | | Insufficient data to meta-analyze | | |
| **BNT162b2** | | | | | | |
| Amit et al (2021)[4] | *n.d.* | *n.d.* | *n.d.* | *n.d.* | *n.d.* | *n.d.* |
| Björk et al (2021)[5] | *V:* 43<br>*UV:* 4,155 | 0.87% | 0.00% | -29%<br>(-1,975%, 92%) | 0.87%<br>(0.58%, 1.15%) | 115<br>(87, 171) |
| Dagan et al (2021)[6] | *V:* 55<br>*UV:* 325 | 1.54% | 3.64% | -136%<br>(-1,088%, 53%) | -2.1%<br>(-7.22%, 3.03%) | -48<br>(33, -14) |
| Fabiani et al (2021)[7] | *n.d.* | *n.d.* | *n.d.* | *n.d.* | *n.d.* | *n.d.* |
| Guijarro et al (2021)[8] | *n.d.* | *n.d.* | *n.d.* | *n.d.* | *n.d.* | *n.d.* |
| Haas et al (2021)[9] | *V:* 6,266<br>*UV:* 109,876 | 0.65% | 2.20% | -238%<br>(-305%, -183%) | -1.55%<br>(-1.92%, -1.19%) | -64<br>(-84, -52) |
| Jones et al (2021)[10] | *n.d.* | *n.d.* | *n.d.* | *n.d.* | *n.d.* | *n.d.* |
| Khan & Mahmud (2021)[15] | *n.d.* | *n.d.* | *n.d.* | *n.d.* | *n.d.* | *n.d.* |
| Menni et al (2021)[19] | *n.d.* | *n.d.* | *n.d.* | *n.d.* | *n.d.* | *n.d.* |
| Monge et al (2021)[11] | *n.d.* | *n.d.* | *n.d.* | *n.d.* | *n.d.* | *n.d.* |
| Moustsen-Helms et al (2021)[12] | *n.d.* | *n.d.* | *n.d.* | *n.d.* | *n.d.* | *n.d.* |
| Polack et al (2020)[13] | *i.d.* | *i.d.* | *i.d.* | *i.d.* | *i.d.* | *i.d.* |
| Pritchard et al (2021)[20] | *n.d.* | *n.d.* | *n.d.* | *n.d.* | *n.d.* | *n.d.* |
| Tang et al (2021)[14] | *n.d.* | *n.d.* | *n.d.* | *n.d.* | *n.d.* | *n.d.* |
| Vasileiou et al (2021)[22] | *n.d.* | *n.d.* | *n.d.* | *n.d.* | *n.d.* | *n.d.* |
| ***Vaccine effect*** | Insufficient data to meta-analyze | | | Insufficient data to meta-analyze | | |
| **mRNA-1273** | | | | | | |
| Baden et al (2021)[23] | *V:* 11<br>*UV:* 185 | 0.54% | 0.00% | -417%<br>(-11,920%, 78%) | 0.54%<br>(-0.52%, 1.60%) | 185<br>(63, -194) |
| Khan & Mahmud (2021)[15] | *n.d.* | *n.d.* | *n.d.* | *n.d.* | *n.d.* | *n.d.* |
| ***Vaccine effect*** | Insufficient data to meta-analyze | | | Insufficient data to meta-analyze | | |
| **ChAdOx1 nCoV-19** | | | | | | |
| Emary et al (2021)[24]‡ | *i.d.* | *i.d.* | *i.d.* | *i.d.* | *i.d.* | *i.d.* |
| Madhi et al (2021)[25] | *n.d.* | *n.d.* | *n.d.* | *n.d.* | *n.d.* | *n.d.* |
| Menni et al (2021)[19] | *n.d.* | *n.d.* | *n.d.* | *n.d.* | *n.d.* | *n.d.* |
| Pritchard et al (2021)[20] | *n.d.* | *n.d.* | *n.d.* | *n.d.* | *n.d.* | *n.d.* |
| Vasileiou et al (2021)[22] | *n.d.* | *n.d.* | *n.d.* | *n.d.* | *n.d.* | *n.d.* |
| Voysey et al (2021a)[26]‡ | *V:* 30<br>*UV:* 101 | 0.99% | 0.00% | -10%<br>(-2,525%, 95%) | 0.99%<br>(-0.94%, 2.92%) | 101<br>(34, -106) |
| Voysey et al (2021b)[27]‡ | *V:* 84<br>*UV:* 248 | 0.40% | 0.00% | 2%<br>(-2,274%, 96%) | 0.40%<br>(-0.39%, 1.19%) | 248<br>(84, -259) |
| ***Vaccine effect*** | Insufficient data to meta-analyze | | | Insufficient data to meta-analyze | | |
| **Ad26.COV2.S** | | | | | | |
| Corchado-Garcia et al (2021)[28] | *V:* 13<br>*UV:* 130 | 0.77% | 0.00% | -212%<br>(-7,199%, 87%) | 0.77%<br>(-0.73%, 2.27%) | 130<br>(44, -136) |
| Sadoff et al (2021)[29] | *V:* 117<br>*UV:* 351 | 1.42% | 0.00% | 73%<br>(-387%, 98%) | 1.42%<br>(0.18%, 2.66%) | 70<br>(38, 541) |
| ***Vaccine effect*** | Insufficient data to meta-analyze | | | Insufficient data to meta-analyze | | |
| **Gam-COVID-Vac** | | | | | | |
| Logunov et al (2021)[30] | *i.d.* | *i.d.* | *i.d.* | *i.d.* | *i.d.* | *i.d.* |
| **WIV04** | | | | | | |
| Kaabi et al (2021)[31] | *i.d.* | *i.d.* | *i.d.* | *i.d.* | *i.d.* | *i.d.* |

| | | | | | | |
|---|---|---|---|---|---|---|
| **HB02** | | | | | | |
| Kaabi et al (2021)[31] | *i.d.* | *i.d.* | *i.d.* | *i.d.* | *i.d.* | *i.d.* |
| **NVX-CoV2373** | | | | | | |
| Shinde et al (2021)[32] | *n.d.* | *n.d.* | *n.d.* | *n.d.* | *n.d.* | *n.d.* |
| **Mixed vaccines** | | | | | | |
| Glampson et al (2021)[17], BNT162b2 and ChAdOx1 nCoV-19 | *n.d.* | *n.d.* | *n.d.* | *n.d.* | *n.d.* | *n.d.* |
| Lumley et al (2021)[18], BNT162b2 and ChAdOx1 nCoV-19 | *n.d.* | *n.d.* | *n.d.* | *n.d.* | *n.d.* | *n.d.* |
| Shrotri et al (2021)[21], BNT162b2 and ChAdOx1 nCoV-19 | *n.d.* | *n.d.* | *n.d.* | *n.d.* | *n.d.* | *n.d.* |
| Swift et al (2021)[16], BNT162b2 and mRNA-1273 | *n.d.* | *n.d.* | *n.d.* | *n.d.* | *n.d.* | *n.d.* |
| ***Vaccine effect*** | Insufficient data to meta-analyze | | | Insufficient data to meta-analyze | | |

'Among infected persons' means the denominators used the number of infections in the vaccinated vs. unvaccinated groups. V=vaccinated, UV=unvaccinated. Overall vaccine effect=meta-analysis of all effects across studies. Vaccine effect=meta-analysis of all effects for a given vaccine. RRR=relative risk reduction (1 – relative risk). ARR=absolute risk reduction. NNV=number needed to vaccinate. 95% CI=95% confidence interval. Brackets are the 95% confidence intervals (lower limit, upper limit). *Note:* displayed risks, RRRs, and ARRs are rounded to two decimal places but were calculated from unrounded numbers. *n.d.*=no data. *i.d.*=insufficient data (i.e., there were no cases in the vaccinated *and* unvaccinated groups).
†Baseline risk is defined as the risk of death from SARS-CoV-2 in the unvaccinated population.
‡Breakthrough risk is the risk of death from SARS-CoV-2 post-vaccination.

**Table S8.** Characteristics and meta-analysis of studies of SARS-CoV-2 vaccines on transmission risk among infected persons

| | | | | | Among infected persons | | | |
|---|---|---|---|---|---|---|---|---|
| **Study and design** | **Vaccine(s)** | **Index case sample size†** | **Contact sample size‡** | **Contact population (vaccination status)§** | **Transmission risks⁑** | **RRR (95% CI)** | **ARR (95% CI)** | **NNV (95% CI)** |
| ***Overall vaccine effect*** | --- | *V:* 27,913<br>*UV:* 516,844<br>*Unclassified:* 12,263 | *V index cases:* 49,328<br>*UV index cases:* 1,294,372 | Closest contacts | *V:* 15.44% (10.16%, 20.73%)<br>$I^2$ = 99.57%<br>Q ($df$ = 9) = 4,229.32<br>$p < 0.0001$<br><br>*UV:* 25.89% (16.14%, 35.64%)<br>$I^2$ = 99.99%<br>Q ($df$ = 7) = 61,230.81<br>$p < 0.0001$ | 40% (29%, 49%)<br>$I^2$ = 97.39%<br>Q ($df$ = 9) = 171.39<br>$p < 0.0001$ | 10.96% (4.98%, 16.95%)<br>$I^2$ = 99.60%<br>Q ($df$ = 9) = 2,365.49<br>$p < 0.0001$ | 9 (6, 20) |
| ***Overall vaccine effect (Large studies only)⸸*** | --- | *V:* 27,848<br>*UV:* 516,581<br>*Unclassified:* 12,263 | *V index cases:* 49,216<br>*UV index cases:* 1,293,631 | Closest contacts | *V:* 14.35% (8.39%, 20.31%)<br>$I^2$ = 99.69%<br>Q ($df$ = 7) = 4,226.74<br>$p < 0.0001$<br><br>*UV:* 23.91% (12.00%, 35.83%)<br>$I^2$ = 99.99%<br>Q ($df$ = 5) = 61,005.87<br>$p < 0.0001$ | 41% (31%, 50%)<br>$I^2$ = 97.75%<br>Q ($df$ = 7) = 166.06<br>$p < 0.0001$ | 11.04% (4.61%, 17.47%)<br>$I^2$ = 99.68%<br>Q ($df$ = 7) = 2,358.74<br>$p < 0.0001$ | 9 (6, 22) |
| de Gier et al (2021)[49]<br><br>Period: 1-Feb-2021 to 27-May-2021<br>Country: Netherlands. | ChAdOx1 nCoV-19 (2 doses), BNT162b2 (2 doses), | *V:* 622<br>*UV:* 110,872 | *V index cases:* 706<br>*UV index cases:* 139,802 | Household (*overall:* 97% unvaccinated, 1% partly vaccinated, 2% fully vaccinated) | *V:* 11.19%<br>*UV:* 30.81% | 64% (55%, 70%) | 19.62% (17.28%, 21.96%) | 5 (5, 6) |

| | | | | | | | | |
|---|---|---|---|---|---|---|---|---|
| Variant: Primarily Alpha. | Ad26.COV2.S (1 dose), or mRNA-1273 (2 doses) | | *V index cases:* 583 *UV index cases:* 108,041 | Other close contacts (*overall:* 95% unvaccinated, 2% partly vaccinated, 3% fully vaccinated) | *V:* 8.58% *UV:* 10.55% | 19% (-6%, 38%) | 1.97% (-0.31%, 4.25%) | 51 (24, -323) |
| de Gier et al (2021)[50] Period: 9-Aug-2021 to 24-Sept-2021 Country: Netherlands. Variant: Primarily Delta. | ChAdOx1 nCoV-19 (2 doses), BNT162b2 (2 doses), Ad26.COV2.S (1 dose), or mRNA-1273 (2 doses) | *V:* 1,740 *UV:* 2,641 | *V index cases:* 2,373 *UV index cases:* 4,022 | Households (*overall:* 38% unvaccinated, 8% partly vaccinated, 54% fully vaccinated) | *V:* 12.39% *UV:* 17.68% | 30% (20%, 38%) | 5.29% (3.51%, 7.06%) | 19 (14, 28) |
| Eyre et al (2021)[51] Period: 2-Jan-2021 to 2-Aug-2021 Country: UK Variant: Alpha (57%) and Delta (43%). | ChAdOx1 nCoV-19 (2 doses) | *V:* 16,680 *UV:* 41,728 | *V index cases:* 25,422 *UV index cases:* 55,977 | Household (70%), household visitors (10%), events/activities (10%), work/education (10%) (*overall:* 51% unvaccinated, 20% partly vaccinated, 29% fully vaccinated) | *V:* 29.73% *UV:* 49.42% | 40% (39%, 41%) | 19.69% (18.99%, 20.39%) | 5 (5, 5) |
| | BNT162b2 (2 doses) | *V:* 4,699 *UV:* 41,728 | *V index cases:* 7,282 *UV index cases:* 55,977 | | *V:* 23.26% *UV:* 49.42% | 53% (51%, 55%) | 26.16% (25.11%, 27.22%) | 4 (4, 4) |
| Harris et al (2021)[52] Period: 4-Jan-2021 to 28-Feb-2021 Country: UK Variant: Not reported. | ChAdOx1 nCoV-19 (at least 1 dose) | *V:* 1,464 *UV:* 361,340 | *V index cases:* 3,424 *UV index cases:* 960,765 | Household (*overall:* 100% unvaccinated) | *V:* 5.72% *UV:* 10.09% | 43% (35%, 50%) | 4.36% (3.58%, 5.14%) | 23 (19, 28) |
| | BNT162b2 (at least 1 dose) | *V:* 2,643 *UV:* 361,340 | *V index cases:* 5,939 *UV index cases:* 960,765 | | *V:* 6.25% *UV:* 10.09% | 38% (32%, 44%) | 3.84% (3.22%, 4.46%) | 26 (22, 31) |
| Layan et al (2021)[53] Period: 31-Dec-2020 to 26-Apr-2021 Country: Israel Variant: Primarily Alpha. | BNT162b2 (2 doses) | *V:* 15 *UV:* 200 | *V index cases:* 43 *UV index cases:* 641 | Household (*overall:* 82% unvaccinated, 18% fully vaccinated) | *V:* 18.60% *UV:* 40.72% | 54% (14%, 76%) | 22.11% (9.88%, 34.35%) | 5 (3, 10) |
| Martínez-Baz et al (2021)[43] | ChAdOx1 nCoV-19 (2 doses), BNT162b2 | *Total N:* 12,263 | *V index cases:* 3,487 | Close contacts (*overall:* 47% unvaccinated, | *V:* 17.55% *UV:* 24.92% | 30% (24%, 35%) | 7.37% (6.00%, 8.74%) | 14 (11, 17) |

| | | | | | | | | |
|---|---|---|---|---|---|---|---|---|
| Period: Apr 2021 to Aug 2021<br>Country: Spain<br>Variant: Alpha (52%), Delta (40%), and non-Alpha/non-Delta (8%). Variant available for 9,041 contacts of fully vaccinated and unvaccinated index cases. | (2 doses), Ad26.COV2.S (1 dose), or mRNA-1273 (2 doses) | *We analyzed the contacts of the fully vaccinated vs. unvaccinated index cases but the authors report only the total number of index cases which included a small number of partially vaccinated index cases who accounted for 6% of all contacts.* | *UV index cases:* 25,024 | 14% partly vaccinated, 39% fully vaccinated) | | | | |
| Singanayagam et al (2021)[54]<br><br>Period: 25-May-2021 to 15-Sept-2021<br>Country: UK<br>Variant: Delta. | Primarily ChAdOx1 nCoV-19 (2 doses), BNT162b2 (2 doses) | *V:* 50<br>*UV:* 63 | *V index cases:* 69<br>*UV index cases:* 100 | Household (*overall:* 22% unvaccinated, 15% partly vaccinated, 63% fully vaccinated) | *V:* 24.64%<br>*UV:* 23.00% | -7%<br>(-85%, 38%) | -1.64%<br>(-14.73%, 11.45%) | -61<br>(9, -7) |

V=vaccinated, UV=unvaccinated. Overall vaccine effect=meta-analysis of all effects across studies. RRR=relative risk reduction (1 – relative risk). ARR=absolute risk reduction. NNV=number needed to vaccinate. 95% CI=95% confidence interval. Brackets are the 95% confidence intervals (lower limit, upper limit). *Note:* displayed risks, RRRs, and ARRs are rounded to two decimal places but were calculated from unrounded numbers. Further, the ARR is the meta-analytic weighted average of the risk differences within studies, not the difference of the overall averages. Thus, the ARR is not simply the overall unvaccinated risk minus the overall vaccinated risk. Variant refers to the variant the contacts were exposed to. If extractable from the study report, the percentages are the percentages of contacts exposed to the variant included in this analysis.

†Total number of index cases (and their vaccination status) of the exposed contacts included in the analysis. The overall vaccine estimates are the sum of the studies, excluding Martínez-Baz et al (2021)[43] because they report only the total number of index cases. This is reported separately as “unclassified”. The unvaccinated sample sizes for the separate vaccines in Eyre et al (2021)[51] and Harris et al (2021)[52] were the same and thus were only counted once towards the overall total.

‡Total number of contacts exposed to vaccinated vs. unvaccinated index cases included in the analysis. The overall vaccine estimates are the sum of the studies. The contacts of unvaccinated index cases for the separate vaccines in Eyre et al (2021)[51] and Harris et al (2021)[52] were the same and thus were only counted once towards the overall total.

§The population the index cases had contact with and the contacts’ vaccination status. Overall=across the full sample of contacts in the study.

⁑The unvaccinated sample sizes for the separate vaccines in Eyre et al (2021)[51] and Harris et al (2021)[52] were the same and thus were only counted once in the meta-analysis of the unvaccinated transmission risk.

⸸Analysis of the transmission studies with large numbers of index cases. Excluded: Layan et al (2021)[53] (*n*=215) and Singanayagam et al (2021)[54] (*n*=113).

**Table S9.** Excluded studies identified in the systematic review of the studies of SARS-CoV-2 vaccines on transmission risk

| Study | Reason for exclusion |
|---|---|
| Apisarnthanarak et al (2021)[55] | Vaccination status of index cases not reported. |
| Bailly et al (2021)[56] | No index case data reported. |
| Bobdey et al (2021)[57] | Vaccination status of index cases not reported. |
| Braeye et al (2021)[58] | Inadequate reporting. Raw sample sizes and infections in the contacts of the vaccinated vs. unvaccinated index cases not reported. |
| Brinkley-Rubinstein et al (2021)[59] | No index case data reported. |
| Brown et al (2021)[60] | No index case data reported. |
| Carter et al (2021)[61] | Vaccination status of index cases not reported. |
| Hetemäki et al (2021)[62] | One index case. |
| Ioannou et al (2021)[63] | No index case data reported. |
| Jahromi & Al Sheikh (2021)[64] | No index case data reported. |
| Kroidl et al (2021)[65] | Case report. |
| Lv et al (2021)[66] | One index case. |
| McEllistrem et al (2021)[67] | No index case data reported. |
| Munitz et al (2021)[68] | No index case data reported. |
| Prato et al (2021)[69] | All index cases were fully vaccinated. |
| Prunas et al (2021)[70] | Inadequate reporting. Raw sample sizes and infections in the contacts of the vaccinated vs. unvaccinated index cases not reported. |
| Salo et al (2021)[71] | Indirect transmission study.† |
| Shah et al (2021)[72] | Indirect transmission study.† |
| Shitrit et al (2021)[73] | One index case. |
| Siddle et al (2021)[74] | Insufficient sample size. Only one unvaccinated index case. |
| Tene et al (2021)[75] | Inadequate reporting. Authors were unable to determine the number of contacts of the unvaccinated index cases and the number of infections within these contacts. |
| Teran et al (2021)[76] | No index case data reported. |

†Indirect transmission studies examine the number of infections within the contacts (e.g., households) of vaccinated vs. unvaccinated persons but do not directly examine the vaccination status of the index cases and utilize methods to identify the contacts of these index cases to determine if they were infected by the index case

**Table S10.** Estimated effects of SARS-CoV-2 vaccines on transmission risk among infected persons and in the general population

| | | | **Baseline infection risk in the general population⁋** | | | | | | | |
|---|---|---|---|---|---|---|---|---|---|---|
| | Among infected persons | | 3.04%‡ | | 5% | | 10% | | 20% | |
| **Setting** | **ARR† (95% CI)** | **NNV (95% CI)** | **ARR† (95% CI)** | **NNV (95% CI)** | **ARR† (95% CI)** | **NNV (95% CI)** | **ARR† (95% CI)** | **NNV (95% CI)** | **ARR† (95% CI)** | **NNV (95% CI)** |
| Closest contacts | 11.04%⁎ (4.61%, 17.47%) | 9⁎ (6, 22) | 0.2980% (0.1496%, 0.4466%) | 336 (224, 669) | 0.4902% (0.2460%, 0.7345%) | 204 (136, 407) | 0.9803% (0.4920%, 1.4690%) | 102 (68, 203) | 1.9606% (0.9840%, 2.9381%) | 51 (34, 102) |
| Household | 4.9200% (4.0180%, 5.8220%) | 20 (17, 25) | 0.1496% (0.1221%, 0.1770%) | 669 (565, 819) | 0.2460% (0.2009%, 0.2911%) | 407 (344, 498) | 0.4920% (0.4018%, 0.5822%) | 203 (172, 249) | 0.9840% (0.8036%, 1.1644%) | 102 (86, 124) |
| Healthcare | 0.2870% (0.1640%, 0.4510%) | 348 (222, 610) | 0.0087% (0.0050%, 0.0137%) | 11,462 (7,294, 20,058) | 0.0144% (0.0082%, 0.0226%) | 6,969 (4,435, 12,195) | 0.0287% (0.0164%, 0.0451%) | 3,484 (2,217, 6,098) | 0.0574% (0.0328%, 0.0902%) | 1,742 (1,109, 3,049) |
| Work or study places | 0.6150% (0.3280%, 0.9430%) | 163 (106, 305) | 0.0187% (0.0100%, 0.0287%) | 5,349 (3,488, 10,029) | 0.0308% (0.0164%, 0.0471%) | 3,252 (2,121, 6,098) | 0.0615% (0.0328%, 0.0943%) | 1,626 (1,060, 3,049) | 0.1230% (0.0656%, 0.1886%) | 813 (530, 1,524) |
| Meal or gathering | 2.5420% (1.8860%, 3.2390%) | 39 (31, 53) | 0.0773% (0.0573%, 0.0985%) | 1,294 (1,016, 1,744) | 0.1271% (0.0943%, 0.1620%) | 787 (617, 1,060) | 0.2542% (0.1886%, 0.3239%) | 393 (309, 530) | 0.5084% (0.3772%, 0.6478%) | 197 (154, 265) |
| Public places | 0.7790% (0.4510%, 1.0660%) | 128 (94, 222) | 0.0237% (0.0137%, 0.0324%) | 4,223 (3,086, 7,294) | 0.0389% (0.0226%, 0.0533%) | 2,567 (1,876, 4,435) | 0.0779% (0.0451%, 0.1066%) | 1,284 (938, 2,217) | 0.1558% (0.0902%, 0.2132%) | 642 (469, 1,109) |
| Daily conversation | 2.2960% (0.0000%, 4.6330%) | 44 (22, Inf) | 0.0698% (0.0000%, 0.1408%) | 1,433 (710, Inf) | 0.1148% (0.0000%, 0.2316%) | 871 (432, Inf) | 0.2296% (0.0000%, 0.4633%) | 436 (216, Inf) | 0.4592% (0.0000%, 0.9266%) | 218 (108, Inf) |
| Transportation | 0.2870% (0.1230%, 0.4920%) | 348 (203, 813) | 0.0087% (0.0037%, 0.0150%) | 11,462 (6,686, 26,744) | 0.0144% (0.0062%, 0.0246%) | 6,969 (4,065, 16,260) | 0.0287% (0.0123%, 0.0492%) | 3,484 (2,033, 8,130) | 0.0574% (0.0246%, 0.0984%) | 1,742 (1,016, 4,065) |

ARR=absolute risk reduction. NNV=number needed to vaccinate. 95% CI=95% confidence interval. Brackets are the 95% confidence intervals (lower limit, upper limit).
⁋Illustration of how the NNV changes as a function of increasing baseline infection risks. As baseline infection risks in the general population increase, ARRs increase, which lowers the NNV.
⁎The ARR and NNV here is from the meta-analysis of the large transmission studies (Table S8).
†ARRs were estimated for each setting by multiplying the corresponding transmission risk in Table 2 by the RRR (0.41, 41%) from the meta-analysis of the large transmission studies (Table S8). See main text for details.

**Table S11.** Estimated transmission risks of vaccinated people among infected persons and in the general population

| | | | **Baseline infection risk in the general population⁋** | | | | | | | |
|---|---|---|---|---|---|---|---|---|---|---|
| | Among infected persons | | 3.04%‡ | | 5% | | 10% | | 20% | |
| **Setting** | **ARR† (95% CI)** | **Vaccinated transmission risk§ (95% CI)** | **ARR† (95% CI)** | **Vaccinated transmission risk§ (95% CI)** | **ARR† (95% CI)** | **Vaccinated transmission risk§ (95% CI)** | **ARR† (95% CI)** | **Vaccinated transmission risk§ (95% CI)** | **ARR† (95% CI)** | **Vaccinated transmission risk§ (95% CI)** |
| Closest contacts | 11.04%⁎ (4.61%, 17.47%) | 14.35%⁎ (8.39%, 20.31%) | 0.2980% (0.1496%, 0.4466%) | 0.4288% (0.0668%, 0.7912%) | 0.4902% (0.2460%, 0.7345%) | 0.7053% (0.1098%, 1.3013%) | 0.9803% (0.4920%, 1.4690%) | 1.4107% (0.2197%, 2.6027%) | 1.9606% (0.9840%, 2.9381%) | 2.8214% (0.4394%, 5.2054%) |
| Household | 4.9200% (4.0180%, 5.8220%) | 7.0800% (4.8800%, 9.2800%) | 0.1496% (0.1221%, 0.1770%) | 0.2152% (0.1484%, 0.2821%) | 0.2460% (0.2009%, 0.2911%) | 0.3540% (0.2440%, 0.4640%) | 0.4920% (0.4018%, 0.5822%) | 0.7080% (0.4880%, 0.9280%) | 0.9840% (0.8036%, 1.1644%) | 1.4160% (0.9760%, 1.8560%) |
| Healthcare | 0.2870% (0.1640%, 0.4510%) | 0.4130% (0.1130%, 0.8130%) | 0.0087% (0.0050%, 0.0137%) | 0.0126% (0.0034%, 0.0247%) | 0.0144% (0.0082%, 0.0226%) | 0.0207% (0.0056%, 0.0406%) | 0.0287% (0.0164%, 0.0451%) | 0.0413% (0.0113%, 0.0813%) | 0.0574% (0.0328%, 0.0902%) | 0.0826% (0.0226%, 0.1626%) |
| Work or study places | 0.6150% (0.3280%, 0.9430%) | 0.8850% (0.1850%, 1.6850%) | 0.0187% (0.0100%, 0.0287%) | 0.0269% (0.0056%, 0.0512%) | 0.0308% (0.0164%, 0.0471%) | 0.0443% (0.0093%, 0.0842%) | 0.0615% (0.0328%, 0.0943%) | 0.0885% (0.0185%, 0.1685%) | 0.1230% (0.0656%, 0.1886%) | 0.1770% (0.0370%, 0.3370%) |
| Meal or gathering | 2.5420% (1.8860%, 3.2390%) | 3.6580% (2.0580%, 5.3580%) | 0.0773% (0.0573%, 0.0985%) | 0.1112% (0.0626%, 0.1629%) | 0.1271% (0.0943%, 0.1620%) | 0.1829% (0.1029%, 0.2679%) | 0.2542% (0.1886%, 0.3239%) | 0.3658% (0.2058%, 0.5358%) | 0.5084% (0.3772%, 0.6478%) | 0.7316% (0.4116%, 1.0716%) |
| Public places | 0.7790% (0.4510%, 1.0660%) | 1.1210% (0.3210%, 1.8210%) | 0.0237% (0.0137%, 0.0324%) | 0.0341% (0.0098%, 0.0554%) | 0.0389% (0.0226%, 0.0533%) | 0.0561% (0.0161%, 0.0910%) | 0.0779% (0.0451%, 0.1066%) | 0.1121% (0.0321%, 0.1821%) | 0.1558% (0.0902%, 0.2132%) | 0.2242% (0.0642%, 0.3642%) |
| Daily conversation | 2.2960% (0.0000%, 4.6330%) | 3.3040% (-2.2960%, 9.0040%) | 0.0698% (0.0000%, 0.1408%) | 0.1004% (-0.0698%, 0.2737%) | 0.1148% (0.0000%, 0.2316%) | 0.1652% (-0.1148%, 0.4502%) | 0.2296% (0.0000%, 0.4633%) | 0.3304% (-0.2296%, 0.9004%) | 0.4592% (0.0000%, 0.9266%) | 0.6608% (-0.4592%, 1.8008%) |
| Transportation | 0.2870% (0.1230%, 0.4920%) | 0.4130% (0.0130%, 0.9130%) | 0.0087% (0.0037%, 0.0150%) | 0.0126% (0.0004%, 0.0278%) | 0.0144% (0.0062%, 0.0246%) | 0.0207% (0.0006%, 0.0457%) | 0.0287% (0.0123%, 0.0492%) | 0.0413% (0.0013%, 0.0913%) | 0.0574% (0.0246%, 0.0984%) | 0.0826% (0.0026%, 0.1826%) |

ARR=absolute risk reduction. 95% CI=95% confidence interval. Brackets are the 95% confidence intervals (lower limit, upper limit).
⁋Illustration of how the vaccination transmission risk changes as a function of increasing baseline infection risks. As baseline infection risks in the general population increase, baseline transmission risks increase, which increases the vaccination transmission risk.
⁎The ARR here is from the meta-analysis of the large transmission studies (Table S8). The vaccinated transmission risk here is the meta-analytic weighted average of the transmission risk in the vaccinated groups of these studies. Note, the ARR is the meta-analytic weighted average of the risk differences within studies, not the difference of the overall averages. Thus, the ARR is not simply the overall unvaccinated risk (23.91%) minus the overall vaccinated risk (14.35%), or 9.56%.
†ARRs taken from Supplementary Table S10.

§Vaccinated transmission risk is defined as the transmission risk in the vaccinated population. Vaccinated transmission risks were estimated for each setting by subtracting the corresponding ARR (Supplementary Table S10) from the corresponding unvaccinated transmission risk (Table 2 in the main text).

**Table S12.** PRISMA 2020 Checklist

| Section and Topic | Item # | Checklist item | Location where item is reported |
|---|---|---|---|
| **TITLE** | | | |
| Title | 1 | Identify the report as a systematic review. | This study is not a traditional systematic review or meta-analysis. It is an estimation and modelling study which uses meta-analyses to calculate key metrics. |
| **ABSTRACT** | | | |
| Abstract | 2 | See the PRISMA 2020 for Abstracts checklist. | pg. 1 - 2 |
| **INTRODUCTION** | | | |
| Rationale | 3 | Describe the rationale for the review in the context of existing knowledge. | pg. 3 – 5 |
| Objectives | 4 | Provide an explicit statement of the objective(s) or question(s) the review addresses. | pg. 5 |
| **METHODS** | | | |
| Eligibility criteria | 5 | Specify the inclusion and exclusion criteria for the review and how studies were grouped for the syntheses. | pg. 6 |
| Information sources | 6 | Specify all databases, registers, websites, organisations, reference lists and other sources searched or consulted to identify studies. Specify the date when each source was last searched or consulted. | pg. 6 |
| Search strategy | 7 | Present the full search strategies for all databases, registers and websites, including any filters and limits used. | pg. 6 |
| Selection process | 8 | Specify the methods used to decide whether a study met the inclusion criteria of the review, including how many reviewers screened each record and each report retrieved, whether they worked independently, and if applicable, details of automation tools used in the process. | pg. 6 -7 |
| Data collection process | 9 | Specify the methods used to collect data from reports, including how many reviewers collected data from each report, whether they worked independently, any processes for obtaining or confirming data from study investigators, and if applicable, details of automation tools used in the process. | pg. 7 |
| Data items | 10a | List and define all outcomes for which data were sought. Specify whether all results that were compatible with each outcome domain in each study were sought (e.g. for all measures, time points, analyses), and if not, the methods used to decide which results to collect. | pg. 6 - 7 |
| | 10b | List and define all other variables for which data were sought (e.g. participant and intervention characteristics, funding sources). Describe any assumptions made about any missing or unclear information. | pg. 7 |

| Section and Topic | Item # | Checklist item | Location where item is reported |
|---|---|---|---|
| Study risk of bias assessment | 11 | Specify the methods used to assess risk of bias in the included studies, including details of the tool(s) used, how many reviewers assessed each study and whether they worked independently, and if applicable, details of automation tools used in the process. | N/A |
| Effect measures | 12 | Specify for each outcome the effect measure(s) (e.g. risk ratio, mean difference) used in the synthesis or presentation of results. | pg. 7 - 10 |
| Synthesis methods | 13a | Describe the processes used to decide which studies were eligible for each synthesis (e.g. tabulating the study intervention characteristics and comparing against the planned groups for each synthesis (item #5)). | pg. 6 |
| | 13b | Describe any methods required to prepare the data for presentation or synthesis, such as handling of missing summary statistics, or data conversions. | pg. 7 |
| | 13c | Describe any methods used to tabulate or visually display results of individual studies and syntheses. | pg. 7 |
| | 13d | Describe any methods used to synthesize results and provide a rationale for the choice(s). If meta-analysis was performed, describe the model(s), method(s) to identify the presence and extent of statistical heterogeneity, and software package(s) used. | pg. 7 - 8 |
| | 13e | Describe any methods used to explore possible causes of heterogeneity among study results (e.g. subgroup analysis, meta-regression). | N/A |
| | 13f | Describe any sensitivity analyses conducted to assess robustness of the synthesized results. | N/A |
| Reporting bias assessment | 14 | Describe any methods used to assess risk of bias due to missing results in a synthesis (arising from reporting biases). | N/A |
| Certainty assessment | 15 | Describe any methods used to assess certainty (or confidence) in the body of evidence for an outcome. | Detailed in the Discussion in terms of statistical power (pg. 18) |
| **RESULTS** | | | |
| Study selection | 16a | Describe the results of the search and selection process, from the number of records identified in the search to the number of studies included in the review, ideally using a flow diagram. | pg. 10 – 11, Supplementary Figure S1 |
| | 16b | Cite studies that might appear to meet the inclusion criteria, but which were excluded, and explain why they were excluded. | Supplementary Tables S2 and S9 |
| Study characteristics | 17 | Cite each included study and present its characteristics. | Supplementary Tables S1 and S8 |
| Risk of bias in studies | 18 | Present assessments of risk of bias for each included study. | N/A |
| Results of individual studies | 19 | For all outcomes, present, for each study: (a) summary statistics for each group (where appropriate) and (b) an effect estimate and its precision (e.g. confidence/credible interval), ideally using structured tables or plots. | Supplementary Tables S3-S7 and S8 |

| Section and Topic | Item # | Checklist item | Location where item is reported |
|---|---|---|---|
| Results of syntheses | 20a | For each synthesis, briefly summarise the characteristics and risk of bias among contributing studies. | N/A |
| | 20b | Present results of all statistical syntheses conducted. If meta-analysis was done, present for each the summary estimate and its precision (e.g. confidence/credible interval) and measures of statistical heterogeneity. If comparing groups, describe the direction of the effect. | Table 1, Table 2, Supplementary Tables S3-S7, S8, S10, and S11 |
| | 20c | Present results of all investigations of possible causes of heterogeneity among study results. | N/A |
| | 20d | Present results of all sensitivity analyses conducted to assess the robustness of the synthesized results. | N/A |
| Reporting biases | 21 | Present assessments of risk of bias due to missing results (arising from reporting biases) for each synthesis assessed. | N/A |
| Certainty of evidence | 22 | Present assessments of certainty (or confidence) in the body of evidence for each outcome assessed. | Detailed in the Discussion in terms of statistical power (pg. 18) |
| **DISCUSSION** | | | |
| Discussion | 23a | Provide a general interpretation of the results in the context of other evidence. | pg. 14 - 16 |
| | 23b | Discuss any limitations of the evidence included in the review. | pg. 17 |
| | 23c | Discuss any limitations of the review processes used. | pg. 17 |
| | 23d | Discuss implications of the results for practice, policy, and future research. | pg. 16 - 17 |
| **OTHER INFORMATION** | | | |
| Registration and protocol | 24a | Provide registration information for the review, including register name and registration number, or state that the review was not registered. | N/A |
| | 24b | Indicate where the review protocol can be accessed, or state that a protocol was not prepared. | N/A |
| | 24c | Describe and explain any amendments to information provided at registration or in the protocol. | N/A |
| Support | 25 | Describe sources of financial or non-financial support for the review, and the role of the funders or sponsors in the review. | 19 |
| Competing interests | 26 | Declare any competing interests of review authors. | 19 |
| Availability of data, code and other materials | 27 | Report which of the following are publicly available and where they can be found: template data collection forms; data extracted from included studies; data used for all analyses; analytic code; any other materials used in the review. | 19 |
